# Bias as a source of inconsistency in ivermectin trials for COVID-19: A systematic review

**DOI:** 10.1101/2021.08.19.21262304

**Authors:** Ariel Izcovich, Sasha Peiris, Martín Ragusa, Fernando Tortosa, Gabriel Rada, Sylvain Aldighieri, Ludovic Reveiz

**Affiliations:** Incident Management System for the Covid-19 Response. Pan American Health Organization, 525 23^rd^ St, NW, Washington DC20037-2895; Evidence and Intelligence for Action in Health Department. Pan American Health Organization, 525 23^rd^ St, NW, Washington DC20037-2895; Fundación Epistemonikos, Holanda 895, Providencia, Santiago, Chile

**Keywords:** COVID-19, severe acute respiratory syndrome coronavirus 2, Coronavirus Infections, Systematic review, ivermectin, bias

## Abstract

**Background and purpose:** The objective of this systematic review is to summarize the effects of ivermectin for the prevention and treatment of patients with COVID-19 and to assess inconsistencies in results from individual studies with focus on risk of bias due to methodological limitations.

**Evidence review:** We searched the L.OVE platform through July 6, 2021 and included randomized trials (RCTs) comparing ivermectin to standard or other active treatments. We conducted random-effects pairwise meta-analysis, assessed the certainty of evidence using the GRADE approach and performed sensitivity analysis excluding trials with risk of bias.

**Results:** We included 29 RCTs which enrolled 5592 cases. Overall, the certainty of the evidence was very low to low. Compared to standard of care, ivermectin may reduce mortality, may increase symptom resolution or improvement, may increase viral clearance, may reduce infections in exposed individuals and may decrease hospitalizations (Risk difference (RD) 21 fewer per 1000, 95%CI: 35 fewer to 4 more). However, after excluding trials classified as “high risk” or “some concerns” in the risk of bias assessment, most estimates of effect changed substantially: Compared to standard of care, low certainty evidence suggests that ivermectin may not significantly reduce mortality (RD 7 fewer per 1000, 95%CI: 77 fewer to 108 more) nor mechanical ventilation (RD 6 more per 1000, 95%CI: 43 fewer to 86 more), and moderate certainty evidence shows that it probably does not significantly increase symptom resolution or improvement (RD 14 more per 1000, 95%CI: 29 fewer to 71 more) nor viral clearance (RD 12 fewer per 1000, 95%CI: 84 fewer to 76 more). It is uncertain if ivermectin increases or decreases severe adverse events and symptomatic infections in exposed individuals.

**Conclusions and Relevance:** Ivermectin may not improve clinically important outcomes in patients with COVID-19 and its effects as a prophylactic intervention in exposed individuals are uncertain. Previous reports concluding significant benefits associated with ivermectin are based on potentially biased results reported by studies with substantial methodological limitations. Further research is needed.

## Introduction

There is an urgent need to expand the evidence base on interventions for the prevention and treatment of COVID-19, an infection caused by SARS-CoV-2 that has the potential of progression into pneumonia, multi-organ failure and death.[1] The COVID-19 pandemic has seen a rapid increase in the number of studies testing potential therapeutic options, raising concerns about the quality and lack of scientific integrity, and also about the spread of this information, leading to the so-called “infodemic”.[2,3] According to the World Health Organization (WHO) international registry of clinical trials platform (ICTRP),[4] hundreds of potential interventions are being assessed in more than 10,000 clinical trials and observational studies.

Many drugs including ivermectin, were repurposed for the treatment of COVID-19, most often based on biological plausibility, in vitro research, or pathophysiological considerations. Ivermectin is a successful broad-spectrum anti-parasitic, included in WHO essential medicines list used to treat several neglected tropical diseases.[5] It emerged as a potential treatment for COVID-19 in mid-2020, following an *in vitro* study demonstrating its anti-viral properties.[6] Multiple systematic reviews have assessed the benefits and harm of ivermectin for COVID-19 patients with inconsistent findings and conclusions.[7] Although some organizations and groups have argued strongly in favor of implementing ivermectin for treatment and/or prevention of COVID-19,[8] current key clinical practice guidelines recommend against its use outside the context of clinical trials.[9–12]

Reasons for these major discrepancies are probably related to different evidence analytical and/or interpretation approaches. Assessing the risk of bias is one of the pillars of any systematic review and has proven to be essential for evidence interpretation in the present pandemic context where results of studies with major methodological limitations have led to erroneous conclusions, waste of resources and patients’ exposure to potentially harmful interventions.[3,13,14] Nevertheless, most available systematic reviews on ivermectin for COVID-19 have not appropriately assessed risk of bias as a potential explanation for inconsistency between trial results. Therefore, this systematic review aims to summarize the best available evidence on ivermectin for prevention and treatment of COVID-19 patients and explore potential explanations for heterogeneity in RCTs results with focus on studies methodological limitations.

## Methods

This systematic review was performed in consistent with Preferred Reporting Items for Systematic Reviews and Meta-Analyses (PRISMA) statement.[15]

### Protocol registration

This systematic review is part of a larger project that aims to conduct multiple systematic reviews for different questions relevant to COVID-19. The protocol stating the shared objectives and methodology of these reviews was published elsewhere.[16]

### Search strategy

We systematically searched in the Living OVerview of Evidence (L.OVE; https://app.iloveevidence.com/covid19) platform, for studies on Ivermectin for COVID-19. L.OVE platform is a system that maps PICO (Patient– Intervention–Comparison–Outcome) questions to a repository developed by Epistemonikos Foundation and is the search platform for the Pan American Health Organization (PAHO) living systematic review of potential therapeutics for COVID-19.^7^ The search terms and databases covered are described on the L.OVE platform methods section available at: https://app.iloveevidence.com/covid19/methods.

The repository that feeds the L.OVE platform was developed and is maintained through the automated and manual screening of multiple databases, trial registries, preprint servers and other sources. The last version of the methods, the total number of sources screened, and a living flow diagram and report of the project is updated regularly on the website. The searches cover the period from inception date of each database. We last searched the platform on July 6, 2021.

There were no restrictions applied to the language or publication status.

### Study selection

Two reviewers (A.I and G.R) working independently and in duplicate, performed study selection, including screening of titles and abstracts and of potentially eligible full-text articles. Reviewers resolved disagreements by discussion.

We included randomized controlled trials (RCTs) that recruited adults with suspected, probable, or confirmed COVID-19, or that were exposed to SARS-COV-2, comparing systemic ivermectin alone or in combination with other drugs, against placebo, standard care or other interventions, and reported on clinical important outcomes (see “Outcomes of interest” below). We included trials regardless of publication status (peer reviewed, in press, or preprint) or language. No restrictions were applied based on severity of COVID-19 illness, setting in which the trial was conducted (e.g. outpatient, inpatient, critical), dose administered or duration of treatment. We excluded studies in which inhaled ivermectin was used as intervention.

### Data extraction

For each eligible trial one reviewer (A.I) extracted data using a standardized, pilot-tested data extraction form. The reviewer collected information on trial characteristics (trial registration, publication status, study status, design), participant characteristics (country, age, sex, comorbidities, and severity), and outcomes of interest. Extracted data was confirmed by a second reviewer (F.T). Discrepancies were resolved through discussion

### Outcomes of interest

We selected clinically important outcomes considering published prioritization exercises performed in the context of different clinical practice guidelines.[9,11] We included all-cause mortality and invasive mechanical ventilation as critical outcomes, and symptom resolution or improvement, hospitalizations, viral clearance, symptomatic infection, and severe adverse events as important outcomes. For symptom resolution or improvement, we considered the proportion of patients with complete resolution of symptoms, or the proportion of patients discharged from hospital or the proportion of patients with significant symptom improvement as reported by investigators. For viral clearance we considered the proportion of patients with negative PCR test. For severe adverse events we used the definition implemented by the investigators.

### Risk of Bias

Two reviewers (A.I and M.R) independently assessed the risk of bias of all included trials using the revised Cochrane Risk of Bias 2.0 tool for randomized trials (RoB 2),[17] focusing on randomization, allocation concealment, blinding, attrition, or other biases relevant to the estimates of effect. In assessing the domain “risk of bias arising from randomization process”, in addition to exploring the balance of baseline prognostic in individual trials, we assessed overall balance by constructing Forest plots. For “mortality” and “mechanical ventilation” outcomes we rated trials at high risk of bias overall if one or more domains were rated as “high risk of bias”, and as “some concerns” if no domains were rated as “high risk of bias” and “Risk of bias arising from randomization process” and/or “Risk of bias due to missing outcome data” and/or “Risk of bias in selection of reported results” domains were classified as “some concerns”. The remaining trials were rated as “low risk of bias”. For other outcomes, we rated trials at high risk of bias overall if one or more domains were rated as “high risk of bias”, as “some concerns” if no domains were rated as “high risk of bias” and one or more domains were rated as “some concerns”, and low risk of bias overall if all domains were rated as “low risk of bias”. Reviewers resolved discrepancies by discussion.

### Data synthesis

We summarized the effect of interventions on selected outcomes using relative risks (RRs) and corresponding 95% confidence intervals (95%CIs). We conducted frequentist random-effects pairwise meta-analyses using the R package “meta” in RStudio Version 1.4.1103.[18] For the primary analysis, we assumed that interventions used in some trials as active comparators (hydroxychloroquine and lopinavir-ritonavir), are not related to significant effects in patients with COVID-19.[9] We considered those interventions as standard of care and performed sensitivity analysis to assess the robustness of results (see subgroup and sensitivity analyses).

### Certainty of the evidence

We assessed the certainty of evidence using the grading of recommendations assessment, development, and evaluation (GRADE) approach.[19] Two methodologists with experience in using GRADE rated each domain for each comparison separately and resolved discrepancies by consensus. We rated the certainty for each comparison and outcome as high, moderate, low, or very low, based on considerations of risk of bias, inconsistency, indirectness, publication bias, and imprecision. We made judgments of imprecision using a minimally contextualised approach with the null effect as a threshold. This minimally contextualised approach considers whether the 95%CI includes the null effect, or, when the point estimate is close to the null effect, whether the 95%CI lies within the boundaries of small but important benefit and harm.[20,21] To define severe or very severe imprecision we considered if the 95%CI included not only the null effect, but significant benefits and harms. We used MAGIC authoring and publication platform (https://app.magicapp.org/) to generate the tables summarizing our findings. We calculated the absolute risks and risk differences from the RRs (and their 95%CIs) and the median risk in the control groups of studies reporting on severe patients for “mortality” and “mechanical ventilation” outcomes. For the remaining outcomes we used RRs (and their 95%CIs) and the median risk in the control groups of all analysed trials.

To communicate our findings and conclusions using statements we followed published guidance.[22]

### Subgroup and sensitivity analyses

To assess if overall estimates of effects could be influenced by trials reporting potentially biased results, we performed sensitivity analysis excluding trials categorized as “high risk of bias” and “some concerns”. We expected smaller effects after excluding those trials. In addition, as there is high certainty evidence on the lack of efficacy of some interventions for the treatment of patients with COVID-19 such as hydroxychloroquine and Lopinavir-Ritonavir,[9] for the primary analysis, we considered those interventions as a part of the standard of care. However, we performed sensitivity analyses excluding trials in which hydroxychloroquine or Lopinavir-Ritonavir were used as comparators. We performed subgroup analysis based on intervention implemented and baseline disease severity, we expected larger effects in trials in which ivermectin was implemented in combination with other interventions and in patients with less severe disease.

### Update of this systematic review

An artificial intelligence algorithm deployed in the COVID-19 topic of the L.OVE platform (https://app.iloveevidence.com/covid19) will provide instant notification of articles with a high likelihood of eligibility. These will be screened by paired reviewers iteratively who will also conduct data extraction and updates of the estimates of effects and certainty of the evidence. We will consider resubmission to a journal if there is a substantial modification of the effect estimate or certainty of the evidence for ivermectin, at the discretion of the reviewer team.

## Results

The search strategy identified 680 potentially eligible records, of which 29 RCTs (reported in 78 references) were included. We identified two additional studies which we decided not to include. One was reported as a cluster randomized trial but methods and results were poorly reported and not consistent with a RCT.[23] The other was mentioned in a published review[24] but we were unable to obtain the full text.[25] We intended to contact the authors of these and other three included studies[26,27,28] for further methodological details by email, but only one responded.[28] On July 14, 2021, one of the included studies was retracted from the preprint server due to research misconduct concerns that are being investigated.[29] As the primary aim of our review was to assess the influence of potentially biased results on ivermectin’s effects interpretation, we decided not to exclude it. The selection process is described by the PRISMA flow diagram in S1 Figure. The list of excluded studies is available upon request.

### Trial characteristics

There was a total of 5592 patients from 29 RCTs,[26–54] in which ivermectin was compared against standard of care or other treatments (S1 Table). Twenty trials were published in peer reviewed journals and nine were only published as preprints. One trial reported the results of three different cohorts, one of severe patients, one of mild patients and one of exposed persons, we therefore analyzed each cohort as a different trial.[29] The sample size ranged from 24 to 1342, with 2830 assigned to Ivermectin and 2483 assigned to control. Eighteen trials included patients with mild to moderate COVID-19,[26,28–30,32–45] three studies included patients with severe to critical COVID-19,[29,46,47] six studies included patients with mild to critical disease,[27,48,49,50,51,52] and four studies included non-infected patients exposed to SARS-COV-2.[29,31,53,54]

Ivermectin administered dose varied from 12 mg once to 400 μgm/kg once a day for 4 days. Ivermectin alone was used in most trials but five in which the intervention implemented was a combination of ivermectin with doxycycline,[26,32,34,48] or iota-Carrageenan.[54] Comparator was standard of care with or without placebo in most trials. Active comparators included hydroxychloroquine or cloroquine,[29,47] hydroxychloroquine plus azythomicin,[32] lopinavirritnonavir[50] and vitamin C.[31]

### Risk of Bias

The risk of bias assessment of the 29 included trials is summarized in table 1. For mortality and mechanical ventilation, our assessment resulted in high risk of bias for four RCT (including the study retracted due to misconduct concerns),[29] some concerns for two and low risk of bias for seven RCTs. For all remaining outcomes, our assessment resulted in high risk of bias for thirteen RCTs, some concerns for nine and low risk of bias for seven RCTs. Most trials did not provide enough information to assess significant baseline differences between arms. Overall assessment of baseline prognostic factors suggested that ischemic heart disease was significantly less frequent in patients assigned to ivermectin (S2 Figure). A detailed description of the trials’ methodological limitations is provided in a supplementary table (S2 Table).

**Table 1.** Risk of bias of included trials.

| Study | Risk-of-bias arising from randomization process | Risk-of-bias due to deviations from the intended interventions | Risk-of-bias due to missing outcome data | Risk-of-bias in measurement of the outcome | Risk-of-bias in selection of the reported result | Overall Risk-of-bias judgement |  |
| --- | --- | --- | --- | --- | --- | --- | --- |
|  |  |  |  |  |  | Mortality and Invasive mechanical ventilation | Symptom resolution or improvement, hospitalization, infection, viral clearance and adverse events |
| Shouman et al<sup>53</sup> | High | Some Concerns | Low | Some Concerns | Low | - | High |
| Chowdhury et al<sup>32</sup> | High | Some Concerns | Low | Some Concerns | Low | - | High |
| Podder et al<sup>33</sup> | High | Some Concerns | Low | Some Concerns | Low | - | High |
| Hashim et al<sup>48</sup> | High | Some Concerns | Low | Some Concerns | Low | High | High |
| Elgazzar et al<sup>29</sup> | High | Some Concerns | Low | Some Concerns | Low | High | High |
| Krolewiecki et al<sup>35</sup> | Low | Some Concerns | Low | Some Concerns | Low | Low | Some concerns |
| Niaee et al<sup>49</sup> | High | Some Concerns | Low | Some Concerns | Low | High | High |
| Ahmed et al<sup>26</sup> | High | Low | Low | Low | Some concerns | - | High |
| Chaccour et al<sup>30</sup> | Low | Low | Low | Low | Low | - | Low |
| Chachar et al<sup>36</sup> | Some Concerns | Some Concerns | Low | Some Concerns | Low | - | Some concerns |
| Babalola et al<sup>50</sup> | Low | Some Concerns | Low | Some Concerns | Low | - | Some concerns |
| Kirti et al<sup>37</sup> | Low | Low | Low | Low | Low | Low | Low |
| Chahla et al<sup>54</sup> | High | Some Concerns | Low | Some Concerns | Low | - | High |
| Mohan et al<sup>38</sup> | Low | Low | Low | Low | Low | - | Low |
| Shahbaznejad et al<sup>51</sup> | Low | Low | Low | Low | Low | Low | Low |
| Samaha et al<sup>39</sup> | High | Some Concerns | Low | Some Concerns | Low | - | High |
| Bukhari et al<sup>40</sup> | High | Some Concerns | Low | Some Concerns | Low | - | High |
| Okumus et al<sup>46</sup> | High | Some Concerns | Low | Some Concerns | Low | High | High |
| Beltran et al<sup>27</sup> | Some Concerns | Low | Low | Low | Low | Some concerns | Some concerns |
| López-Medina et al<sup>41</sup> | Low | Low | Low | Low | Low | Low | Low |
| Bermejo Galan et al<sup>47</sup> | Low | Low | Low | Low | Low | Low | Low |
| Pott-Junior et al<sup>52</sup> | Low | Some Concerns | Low | Some Concerns | Low | - | Some concerns |
| Kishoria et al<sup>42</sup> | Low | Some Concerns | Low | Some Concerns | Low | - | Some concerns |
| Seet et al<sup>31</sup> | Low | Some Concerns | Low | Some Concerns | Low | - | High |
| Mahmud et al<sup>33</sup> | Low | Low | Some Concerns | Low | Low | Some concerns | Some concerns |
| Abd-Elsalam et al<sup>43</sup> | Low | Some Concerns | Low | Some Concerns | Low | Low | Some concerns |
| Biber et al<sup>44</sup> | Some Concerns | Low | Some Concerns | Low | Low | - | Some concerns |
| Faisal et al<sup>45</sup> | High | Some Concerns | Low | Some Concerns | Low | - | High |
| Vallejos et al<sup>28</sup> | Low | Low | Low | Low | Low | Low | Low |

### Effects on assessed outcomes

Table 2. and Figure 1. provide a summary of finding for all assessed outcomes.

**Table 2.**
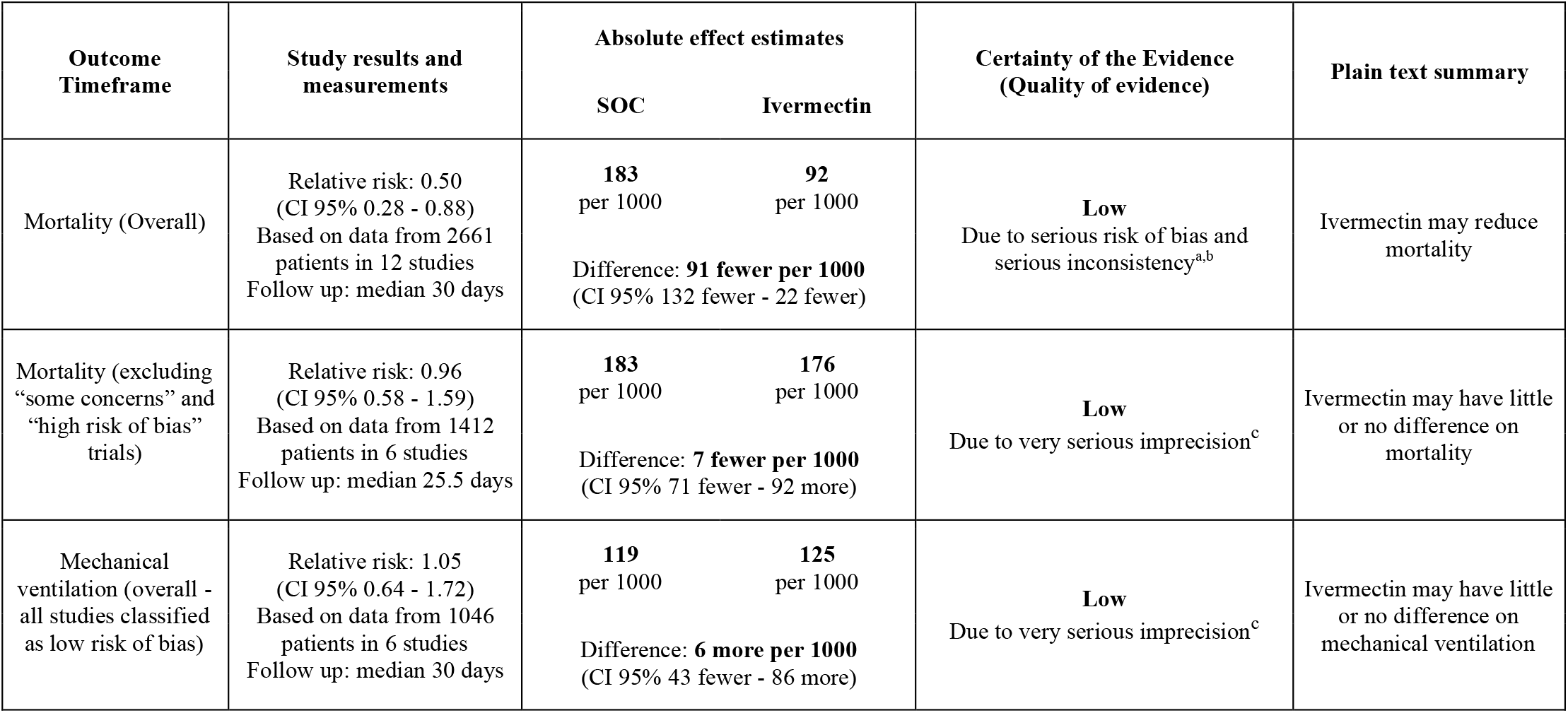
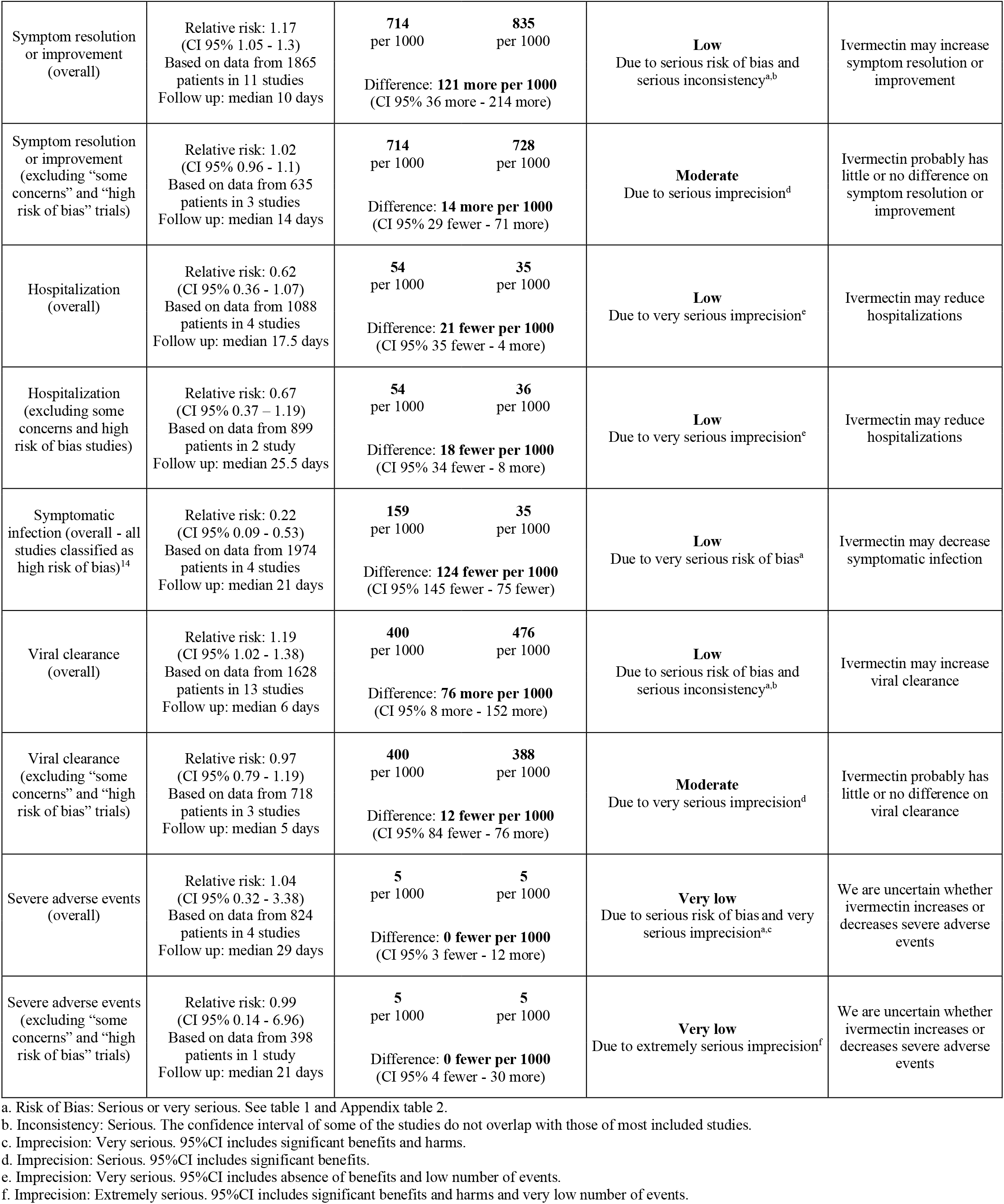
Summary of findings table.

**Figure 1.**
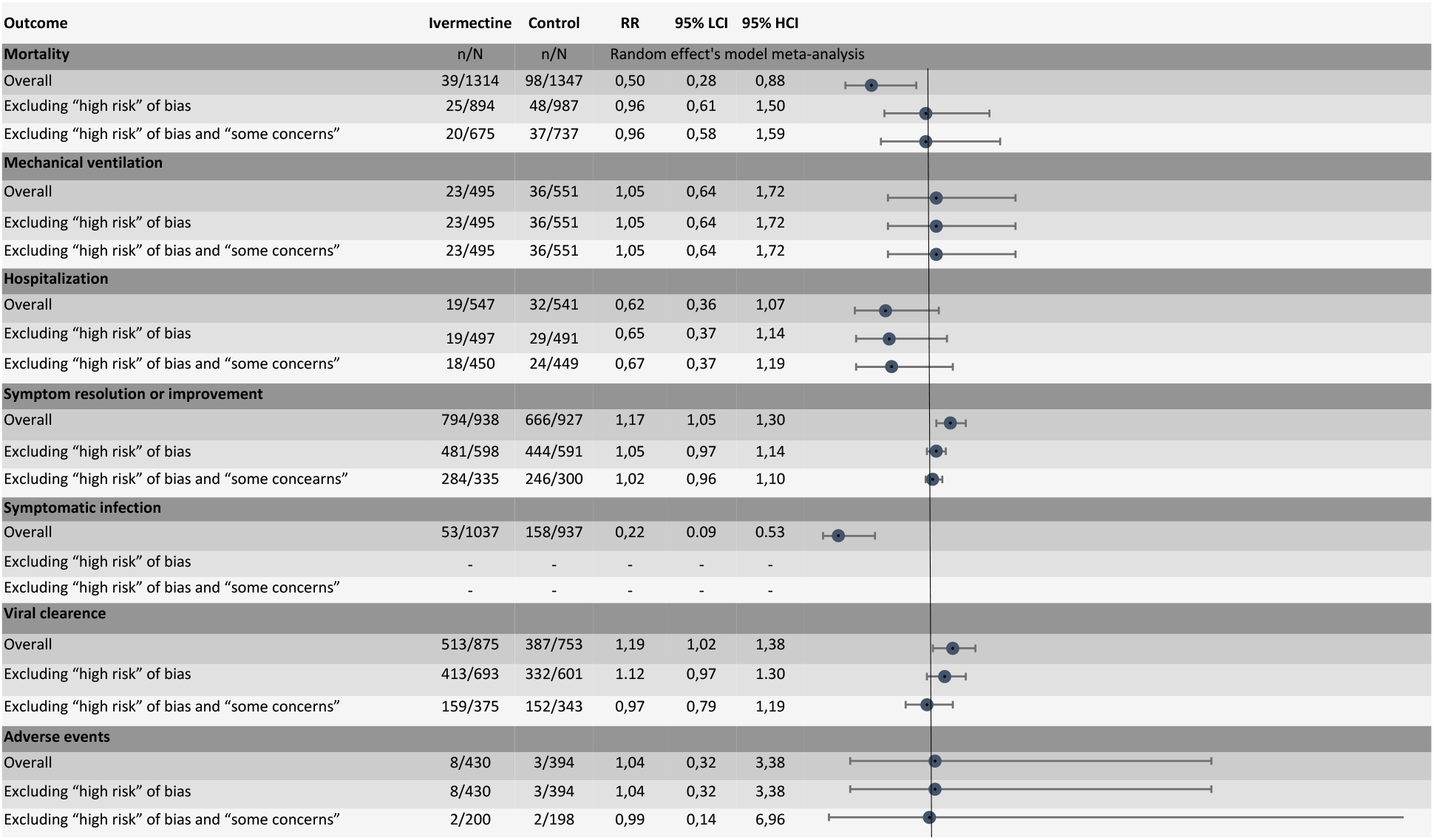
Results of primary analysis and sensitivity analysis excluding trials with significant methodological limitations.

### Mortality

Twelve trials with 2661 patients reported on mortality.[27,28,29,34,37,41,43,46–49,51] Ivermectin may reduce mortality (RR 0.50, 95% CI:0.28 to 0.88; RD 91 fewer per 1,000 participants, 95% CI: 132 fewer to 22 fewer). The certainty of the evidence was low because of serious risk of bias and serious inconsistency (I^2^ 48%). Sensitivity analysis excluding six trials classified as “some concerns” or “high risk of bias” showed that ivermectin may not significantly reduce mortality (RR 0.96, 95% CI:0.58 to 1.59; RD 7 fewer per 1,000 participants, 95% CI: 71 fewer to 92 more) (Figure 2).

**Figure 2.**
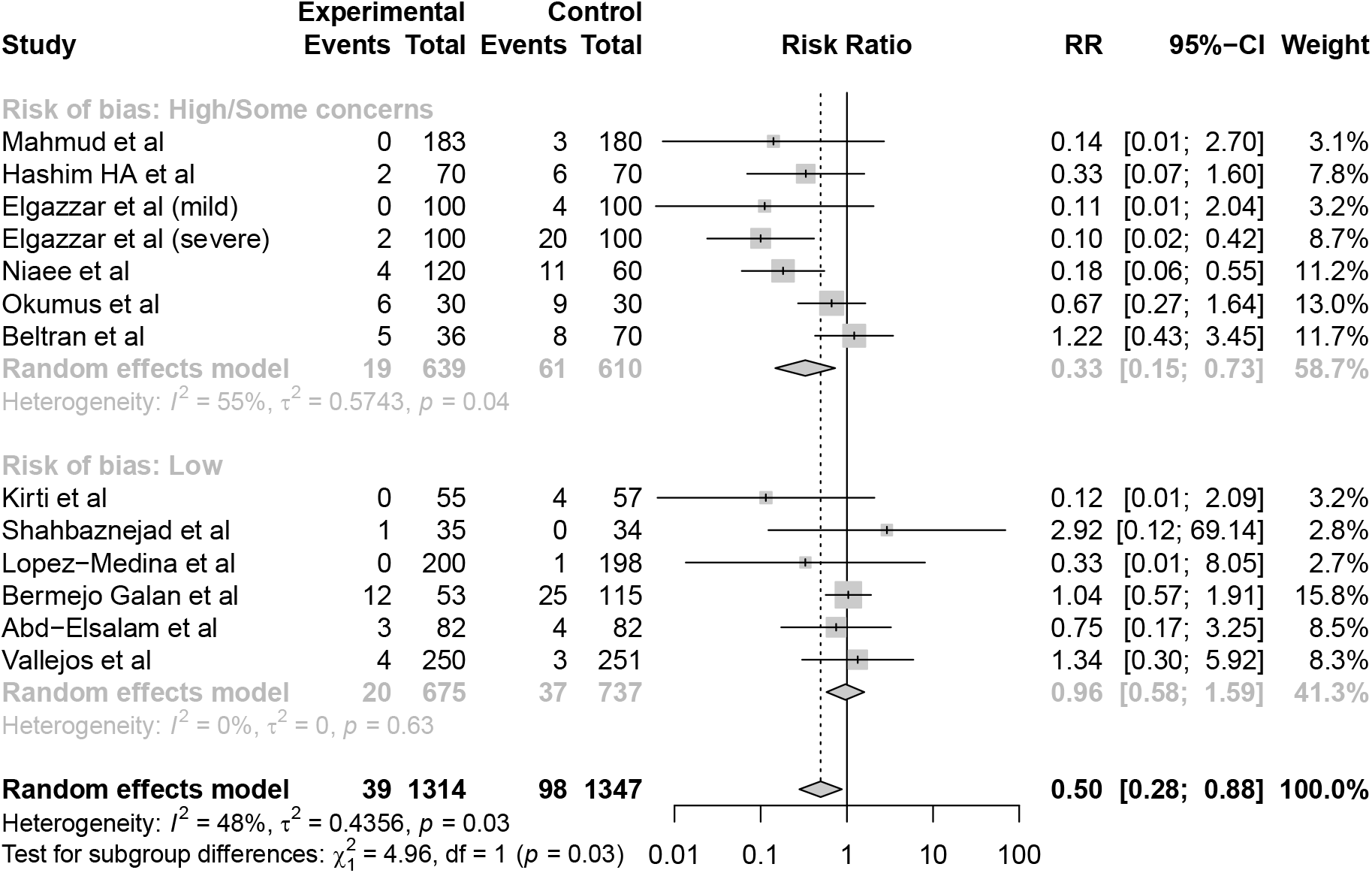
Comparison: ivermectin vs. Standard of care; Outcome: mortality; Analysis: subgroups by risk of bias classification.

### Mechanical ventilation

Six trials with 1046 patients, classified as “low risk of bias”, reported on mechanical ventilation.[28,35,37,43,47,51] Ivermectin may not reduce the requirement of mechanical ventilation (RR 1.05, 95% CI:0.64 to 1.72; RD 6 more per 1,000 participants, 95% CI: 43 fewer to 86 more) (S3 Figure in S1 Appendix). The certainty of the evidence was low because of very serious imprecision.

### Symptom resolution or improvement

Eleven trials with 1865 patients reported on symptom resolution or improvement.[27,29,32,34,36–38,41,42,45,46] Ivermectin may increase symptom resolution or improvement (RR 1.17, 95% CI:1.05 to 1.30); RD 121 more per 1,000 participants, 95% CI: 36 more to 214 more). The certainty of the evidence was low because of serious risk of bias and serious inconsistency (I^2^ 79%). Sensitivity analysis excluding eight trials classified as “some concerns” or “high risk of bias” showed that ivermectin probably does not significantly increase symptom resolution or improvement (RR 1.02, 95% CI:0.96 to 1.10; RD 14 more per 1,000 participants, 95% CI: 29 fewer to 71 more) (S4 Figure in S1 Appendix).

### Hospitalization

Four trials with 1088 patients reported on hospitalization.[28,39,41,44] Ivermectin may decrease hospitalizations (RR 0.62, 95% CI:0.36 to 1.07); RD 21 fewer per 1,000 participants, 95% CI: 35 fewer to 4 more). The certainty of the evidence was low because of very serious imprecision. Sensitivity analysis excluding two trials classified as “some concerns” or “high risk of bias” showed that ivermectin may decrease hospitalizations (RR 0.67, 95% CI:0.37 to 1.19); RD 18 fewer per 1,000 participants, 95% CI: 34 fewer to 10 more) (S5 Figure in S1 Appendix).

### Symptomatic infection in exposed persons

Four trials including 1974 patients, classified as “high risk of bias”, reported on symptomatic infection.[29,31,53,54] Ivermectin may reduce symptomatic infection (RR 0.22, 95% CI:0.09 to 0.53); RD 124 fewer per 1,000 participants, 95% CI: 145 fewer to 75 fewer) (S6 Figure in S1 Appendix). The certainty of the evidence was low because of very serious risk of bias.

### Viral clearance

Thirteen trials with 1628 patients reported on viral clearance.[26,28,32,33,34,37,38,40,42,44,46,50,52] Ivermectin may increase viral clearance (RR 1.19, 95% CI: 1.02 to 1.38); RD 76 more per 1,000 participants, 95% CI: 8 more to 152 more). The certainty of the evidence was low because of serious risk of bias and serious inconsistency (I^2^ 56%). Sensitivity analysis excluding ten trials classified as “some concerns” or “high risk of bias” showed that ivermectin probably does not significantly increase viral clearance (RR 0.97, 95% CI: 0.79 to 1.19); RD 12 fewer per 1,000 participants, 95% CI: 84 fewer to 76 more) (S7 Figure in S1 Appendix).

### Severe Adverse events

Four trials with 824 patients reported on severe adverse events.[34,35,41,52] It is uncertain if Ivermectin increases or decreases severe adverse events (RR 1.04, 95% CI: 0.32 to 3.38); RD 0 more per 1,000 participants, 95% CI: 3 fewer to 12 more). The certainty of the evidence was very low because of serious risk of bias and very serious imprecision. Sensitivity analysis excluding ten trials classified as “some concerns” or “high risk of bias” showed that it is uncertain if ivermectin increases or decreases severe adverse events (RR 0.99, 95% CI: 0.14 to 6.96); RD 0 more per 1,000 participants, 95% CI: 4 fewer to 30 more) (S8 Figure in S1 Appendix).

### Additional analysis

Subgroup and sensitivity analysis did not suggest differential effects according to baseline disease severity, or when ivermectin was administered in combination with other interventions, or when it was compared against hydroxychloroquine or lopinavir-ritonavir, or when different outcome measurements time frames were used (S9 to S30 Figures in S2 Appendix). Visual inspection of the funnel plot for mortality suggested possible publication bias, (S31 Figure in S2 Appendix) however egger’s test was not statistically significant (p=0.13). Visual inspection of funnel plots for symptom resolution or improvement and viral clearance did not suggest publication bias, egger’s test results p=0.48 and p=0.25 respectively (S32 and S33 Figures in S2 Appendix).

## Discussion

This systematic review and meta-analysis provide a comprehensive overview of the available evidence on ivermectin for prevention and treatment of COVID-19. Overall, the body of evidence suggests that ivermectin may reduce mortality, may increase symptom resolution or improvement, may decrease hospitalizations, may increase viral clearance, and may decrease symptomatic infection in exposed individuals. However most trials have serious methodological limitations including lack of allocation concealment and lack of blinding, and reported results varied significantly from striking benefits to null effects. GRADE assessment resulted in low or very low certainty of the evidence for all the outcomes, due to risk of bias, inconsistency, and imprecision. Visual inspection of funnel plot constructed for mortality outcome suggest possible publication bias which rises additional concerns about the certainty of the evidence on ivermectin’s effects.

After excluding trials with significant methodological limitations inconsistency disappeared and results changed substantially. We found low certainty, due to imprecision, that ivermectin may not significantly reduce mortality, nor reduce invasive mechanical ventilation, and moderate certainty evidence that ivermectin probably does not significantly increase viral clearance or symptom resolution or improvement. Regarding hospitalizations, results did not change significantly suggesting that ivermectin may modestly reduce hospitalizations. However, certainty of the evidence remained low due to very serious imprecision. It is uncertain if Ivermectin reduces or increases symptomatic infections in exposed individuals or increases severe adverse events as no trials classified as “low risk of bias” were identified, or the certainty of the evidence was very low.

Our systematic review has several strengths. The search strategy was comprehensive with explicit eligibility criteria, and no restrictions on language or publication status. We used a validated tool for risk of bias assessment and performed a thorough assessment providing details of trial limitations and potential significant imbalances in baseline participant characteristics. We assessed the certainty of the evidence using the GRADE approach and interpreted the results considering absolute rather than relative effects.

Reporting was poor for a significant number of included trials. For risk of bias assessment, we adopted a conservative approach and rated as low risk of bias only those trials for which it was clearly reported that no significant methodological limitations existed. Hence, we may have inappropriately classified some well executed trials as “some concerns” or “high risk of bias” due to their suboptimal reporting methods. Although for some trials we intended to contact the authors for clarification, most did not answer.

Multiple systematic reviews assessed ivermectin for COVID-19.[7] Most of these reviews were already outdated at the time of writing this manuscript.[55] Only four reviews included a substantial proportion of the studies assessed in our review.[56–59] In agreement with our findings, all these reviews concluded that most of the studies assessing ivermectin for COVID-19 have considerable methodological limitations, and two judged the certainty of the evidence as low to very low for all outcomes[56] or not robust enough to justify ivermectin’s use.[57] The authors of one systematic review concluded that ivermectin “may have a role in decreasing mortality in mildly/moderately ill COVID-19 patients” although they graded the certainty on ivermectin’s effect on mortality as very low.[58] Bryant el at. graded the certainty of the evidence as low or very low for all outcomes except mortality for which they report moderate certainty in important mortality reduction. In contrast to our analysis, they reached this conclusion by not downgrading the certainty of the evidence for inconsistency even though they reported there was significant, not fully explained, heterogeneity in studies’ results. In addition, for mortality outcome, they report a sensitivity analysis excluding high risk of bias studies which, in contrast to our findings, did not result in different estimates of effect from the primary analysis. This can be explained by the fact that the authors did not exclude a significant number of studies with important methodological limitations, that they classified as “unclear” risk of bias.[24]

Due to the excessive amount of rapidly published research on COVID-19, often referred to as an “infodemic”,[2,3] the scientific community has already faced a similar scenario to the one described for ivermectin in the present review. Small studies with significant methodological limitations suggested benefits for steroids, lopinavir-ritonavir, interferon β-1a and convalescent plasma among others.[59–62] However, those potential benefits were seldom confirmed and mostly discarded by well-designed adequately powered studies.[63–66] The limitations in the body of evidence on ivermectin for COVID-19 does not allow to reach firm conclusions, however the results of our analysis highlight that most of current suggested benefits of ivermectin are based on potentially biased estimates reported by studies with significant methodological limitations. Further research is needed to confirm or reject the effects of ivermectin on patient important outcomes.

There is an urgent need for high quality research both in health emergencies and in health relevant priorities in non-emergency settings. Although countries have capacities to conduct trials, and there exist global standards of quality assurance in clinical trials,[67–69] a global coordinating mechanism is needed to streamline and harmonize research findings on an international scale.

## Conclusions

Ivermectin may not improve clinically important outcomes in patients with COVID-19 and its effects as a prophylactic intervention in exposed individuals are uncertain. Previous reports concluding significant benefits associated with ivermectin are based on potentially biased results reported by studies with significant methodological limitations. Further research is needed.

## Supporting information

Primary analysis Forest plots

PRISMA (preferred reporting items for systematic reviews and meta-analyses) flowchart of study inclusions and exclusions

Included randomized controlled trials characteristics

Additional analysis plots

Baseline prognostic factors balance between study arms

## Data Availability

All data is available in the main manuscript and/or supplementary materials

## Acknowledgments

We would like to thank Victoria Stanford for contributing to the writing of the final version of the manuscript.

## Disclaimer

The authors have declared that no competing interests exist. Authors hold sole responsibility for the views expressed in the manuscript, which may not necessarily reflect the opinion or policy of the Pan American Health Organization.

## Supplementary files

**S1 Figure. PRISMA (preferred reporting items for systematic reviews and meta-analyses) flowchart of study inclusions and exclusions**

**S2 Figure. Baseline prognostic factors balance between study arms**

**S1 Table. Included randomized controlled trials characteristics**

**S2 Table. Included trials’ methodological limitations**

**S1 Appendix. Primary analysis Forest plots**

**S2 Appendix. Additional analysis plots**

