## Supplementary material for "Bias as a source of inconsistency in ivermectin trials for COVID-19: A systematic review": Primary analysis Forest plots

**S3 Figure. Comparison: ivermectin vs. Standard of care; Outcome: mechanical ventilation; Analysis: subgroups by risk of bias classification.**

| Study | Experimental Events | Experimental Total | Control Events | Control Total | RR | 95%−CI | Weight |
|---|---|---|---|---|---|---|---|
| **Risk of bias: Low** | | | | | | | |
| Krolewiecki et al | 1 | 20 | 0 | 12 | 1.83 | [0.08; 41.52] | 2.5% |
| Kirti et al | 1 | 55 | 5 | 57 | 0.21 | [0.03; 1.72] | 5.5% |
| Shahbaznejad et al | 2 | 35 | 1 | 34 | 1.94 | [0.18; 20.45] | 4.5% |
| Bermejo Galan et al | 12 | 53 | 24 | 115 | 1.08 | [0.59; 2.00] | 66.2% |
| Abd−Elsalam et al | 3 | 82 | 3 | 82 | 1.00 | [0.21; 4.81] | 10.0% |
| Vallejos et al | 4 | 250 | 3 | 251 | 1.34 | [0.30; 5.92] | 11.2% |
| **Random effects model** | **23** | **495** | **36** | **551** | **1.05** | **[0.64; 1.72]** | **100.0%** |
| Heterogeneity: $I^2 = 0\%$, $\tau^2 = 0$, $p = 0.73$ | | | | | | | |
| **Random effects model** | **23** | **495** | **36** | **551** | **1.05** | **[0.64; 1.72]** | **100.0%** |
| Heterogeneity: $I^2 = 0\%$, $\tau^2 = 0$, $p = 0.73$ | | | | | | | |
| Test for subgroup differences: $\chi^2_0 = 0.00$, df = 0 ($p$ = NA) | | | | | | | |

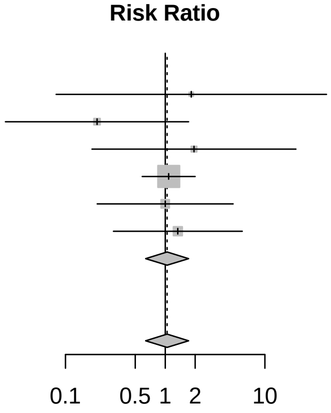


**S4 Figure. Comparison: ivermectin vs. Standard of care; Outcome: symptom resolution or improvement; Analysis: subgroups by risk of bias classification.**

| Study | Experimental Events | Experimental Total | Control Events | Control Total | RR | 95%−CI | Weight |
|---|---|---|---|---|---|---|---|
| **Risk of bias: High/Some concerns** | | | | | | | |
| Chowdhury et al | 50 | 60 | 40 | 56 | 1.17 | [0.95; 1.43] | 8.6% |
| Mahmud et al | 141 | 183 | 113 | 180 | 1.23 | [1.07; 1.41] | 10.2% |
| Elgazzar et al (mild) | 99 | 100 | 74 | 100 | 1.34 | [1.19; 1.51] | 10.7% |
| Elgazzar et al (severe) | 94 | 100 | 50 | 100 | 1.88 | [1.54; 2.30] | 8.5% |
| Chachar et al | 16 | 25 | 15 | 25 | 1.07 | [0.69; 1.65] | 4.1% |
| Okumus et al | 22 | 30 | 16 | 30 | 1.38 | [0.92; 2.05] | 4.5% |
| Beltran et al | 32 | 36 | 64 | 70 | 0.97 | [0.85; 1.11] | 10.2% |
| Kishoria et al | 8 | 19 | 6 | 16 | 1.12 | [0.49; 2.56] | 1.5% |
| Faisal et al | 48 | 50 | 42 | 50 | 1.14 | [1.00; 1.31] | 10.3% |
| **Random effects model** | **510** | **603** | **420** | **627** | **1.24** | **[1.08; 1.43]** | **68.6%** |
| Heterogeneity: $I^2 = 77\%$, $\tau^2 = 0.0290$, $p < 0.01$ | | | | | | | |
| **Risk of bias: Low** | | | | | | | |
| Kirti et al | 46 | 55 | 51 | 57 | 0.93 | [0.81; 1.08] | 10.0% |
| Mohan et al | 74 | 80 | 39 | 45 | 1.07 | [0.94; 1.22] | 10.4% |
| Lopez−Medina et al | 164 | 200 | 156 | 198 | 1.04 | [0.94; 1.15] | 11.1% |
| **Random effects model** | **284** | **335** | **246** | **300** | **1.02** | **[0.96; 1.10]** | **31.4%** |
| Heterogeneity: $I^2 = 0\%$, $\tau^2 = 0$, $p = 0.37$ | | | | | | | |
| **Random effects model** | **794** | **938** | **666** | **927** | **1.17** | **[1.05; 1.30]** | **100.0%** |
| Heterogeneity: $I^2 = 79\%$, $\tau^2 = 0.0244$, $p < 0.01$ | | | | | | | |
| Test for subgroup differences: $\chi^2_1 = 6.07$, df = 1 ($p$ = 0.01) | | | | | | | |

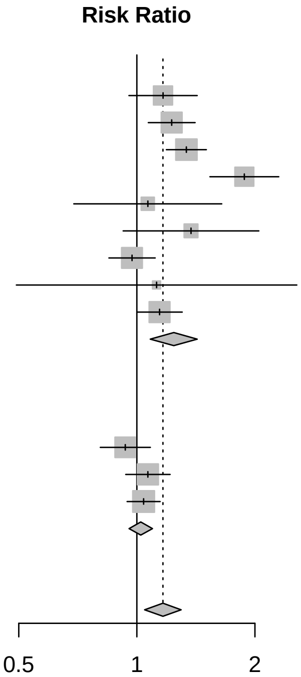


**S5 Figure. Comparison: ivermectin vs. Standard of care; Outcome: hospitalization; Analysis: subgroups by risk of bias classification.**

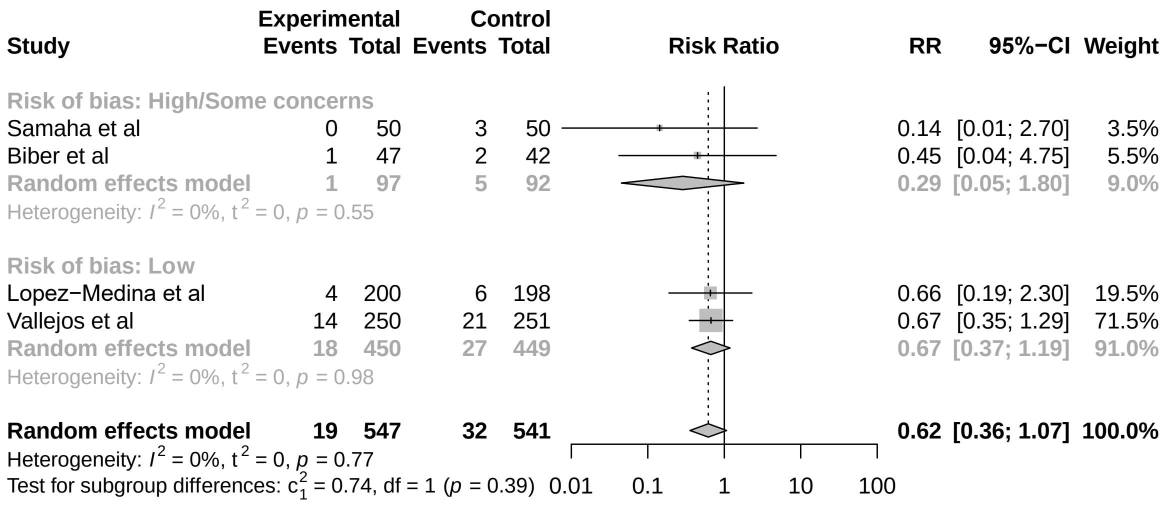


**S6 Figure. Comparison: ivermectin vs. Standard of care; Outcome: symptomatic infection; Analysis: subgroups by risk of bias classification.**

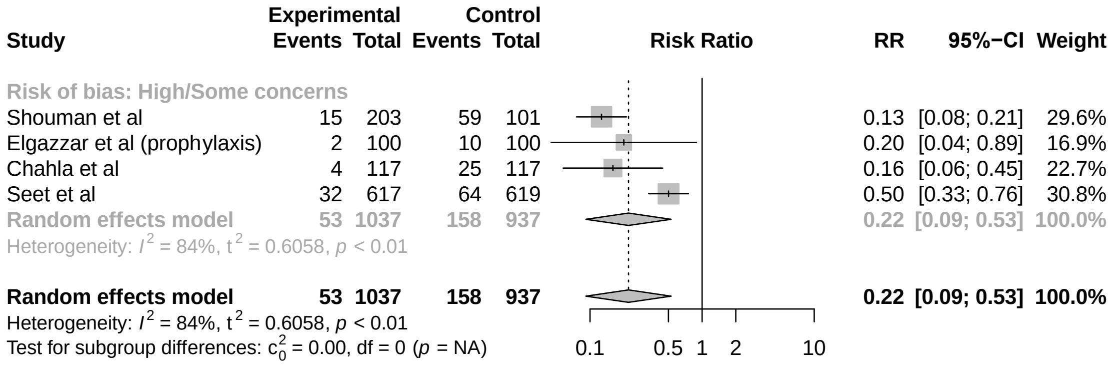


**S7 Figure. Comparison: ivermectin vs. Standard of care; Outcome: viral clearance; Analysis: subgroups by risk of bias classification.**

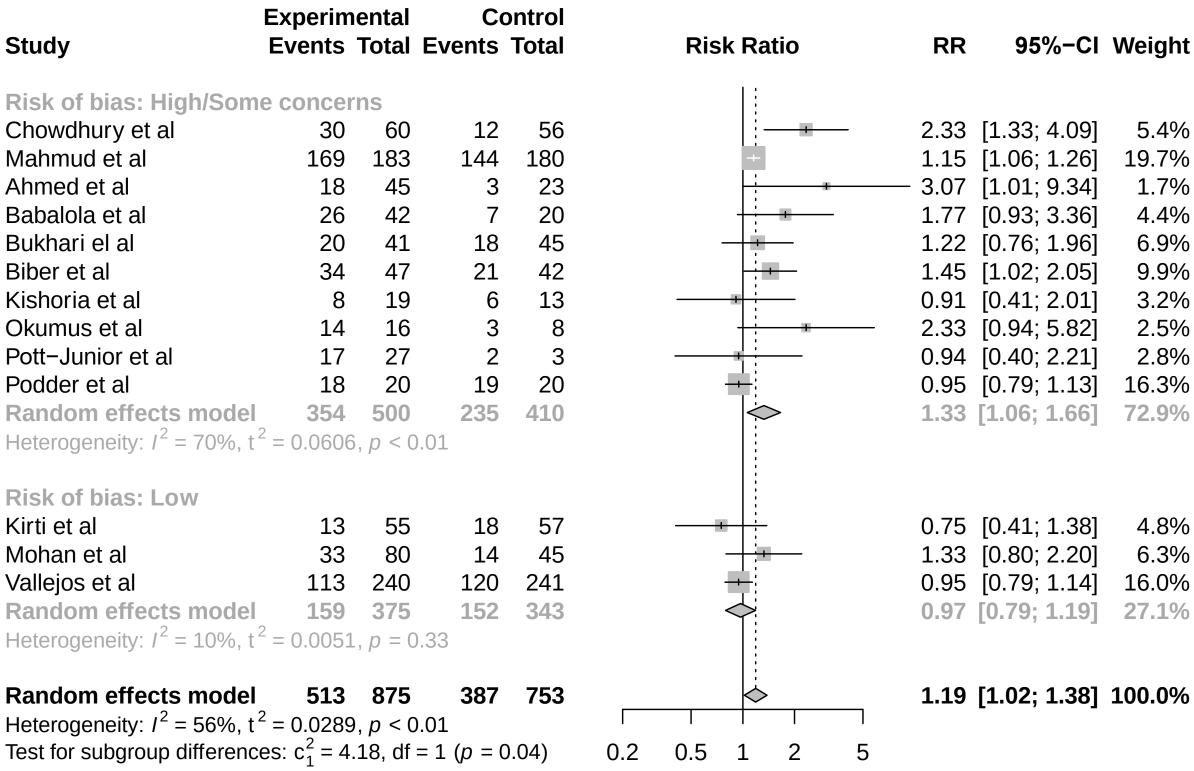


**S8 Figure. Comparison: ivermectin vs. Standard of care; Outcome: adverse events; Analysis: subgroups by risk of bias classification.**

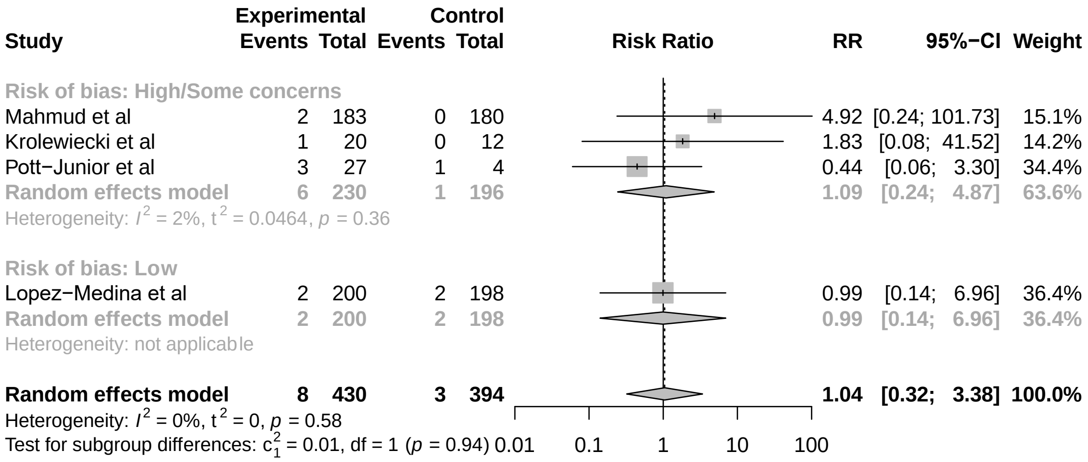
