## Supplementary material for "Bias as a source of inconsistency in ivermectin trials for COVID-19: A systematic review": PRISMA (preferred reporting items for systematic reviews and meta-analyses) flowchart of study inclusions and exclusions

**S1 Figure. PRISMA (preferred reporting items for systematic reviews and meta-analyses) flowchart of study inclusions and exclusions**

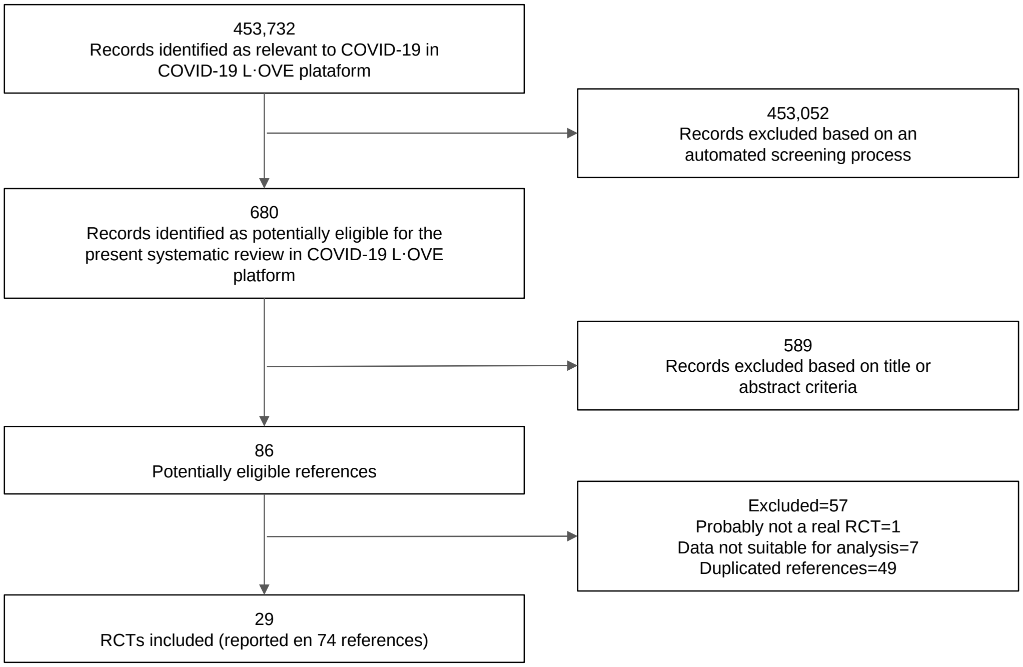
