## Supplementary material for "Bias as a source of inconsistency in ivermectin trials for COVID-19: A systematic review": Included randomized controlled trials characteristics

**S1 Table. Included randomized controlled trials characteristics**

| Study | Publication status | Baseline severity | N intervention group | N control group | Patients' characteristics | Intervention | Control | Standard of care (SOC) implemented | Additional interventions |
|---|---|---|---|---|---|---|---|---|---|
| Shouman et al; 2020 | Peer-reviewed | Patients exposed to COVID-19 (ambulatory) | 228 | 112 | Mean age 38.72 ± 15.94, male 51.3%, HTN 9.5%, DM 7.6%, CKD 1%, asthma 3%, CHD 2.3% | Ivermectin 15 to 24 mg twice (second dose at day 3) | No intervention | NR | NR |
| Chowdhury et al; 2020 | Pre-print | Patients with mild to moderate COVID-19 (ambulatory) | 63 | 62 | Mean age 33.9 ± 14.1, male 72.4% | Ivermectin plus doxycycline 200 µgm/kg single dose + 100 mg BID for 10 days and | HCQ 200 mg daily for 10 days + AZT 500 mg daily for 5 days | NR | NR |
| Podder et al; 2020 | Peer-reviewed | Patients with mild to moderate COVID-19 (ambulatory) | 32 | 30 | Mean age 39.16 ± 12.07, male 71% | Ivermectin 200 µgm/kg once + SOC | SOC | Antipyretics, cough suppressants, and capsule doxycycline (100 mg every 12 hours for seven days) | NR |
| Hashim et al; 2020 | Pre-print | Patients with mild to severe COVID-19 (hospitalized) | 70 | 70 | Mean age 48.7 ± 8.6, male 52% | Ivermectin 200 µgm/kg two or three doses + doxycycline 100 mg twice a day for 5 to 10 days + SOC | SOC | Vitamin C 1000 mg twice/ day and Zinc 75-125 mg/day and Vitamin D3 5000IU/day plus Dexamethasone 6 mg/day or methylprednisolo | Steroids 100%, azithromycin 100%, |

| | | | | | | | | | |
|---|---|---|---|---|---|---|---|---|---|
| | | | | | | | | ne 40mg twice per day, if needed | |
| Mahmud et al; 2020 | Peer-reviewed | Patients with mild to moderate COVID-19 (hospitalized) | 200 | 200 | Mean age 39.6 ± 13.2, male 58.8%, HTN 14%, DM 13%, asthma 5%, CKD 2% | Ivermectin plus doxycycline 12 mg once + 100 mg twice a day for 5 days + SOC | Placebo + SOC | Paracetamol, antihistamines, cough suppressants, vitamins, low molecular weight heparin, remdesivir injection | NR |
| Elgazzar et al (mild);[151] 2020 | Pre-print | Patients with mild to moderate COVID-19 (hospitalized) | 100 | 100 | Mean age 55.2 ± 19.8, male 69.5%, HTN 11.5%, DM 14.5%, COPD %, asthma 5.5%, CHD 4% | Ivermectin 400 µgm/kg once for 4 days + SOC and 100 patients assigned to | HCQ 800 mg for one day and 400 mf for 4 days + SOC | Azithromycin 500mg OD for 6 days, Paracetamol 500mg PRN, vitamin C 1gm OD, Zinc 50 mg OD, Lactoferrin 100mg sachets BID , Acetylcystein 200mg t.d.s & prophylactic or therapeutic anticoagulation if D-dimer > 1000 | NR |
| Elgazzar et al (severe);[151] 2020 | Pre-print | Patients with severe COVID-19 (hospitalized) | 100 | 100 | Mean age 39.6 ± 13.2, male 58.8%, HTN 14%, DM 13%, asthma 5%, CKD 2% | Ivermectin 400 µgm/kg once for 4 days + SOC | HCQ 800 mg for one day and 400 mg for 9 days + SOC | Azithromycin 500mg OD for 6 days, Paracetamol 500mg PRN, | NR |

| | | | | | | | | | |
|---|---|---|---|---|---|---|---|---|---|
| | | | | | | | | vitamin C 1gm OD, Zinc 50 mg OD, Lactoferrin 100mg sachets BID , Acetylcystein 200mg sachets t.d.s , prophylactic or therapeutic anticoagulation if D-dimer > 1000 and systemic steroids | |
| Elgazzar et al (prophylaxis);[151] 2020 | Pre-print | People exposed to SARS-COV-2 infection (ambulatory) | 100 | 100 | Mean age: 57.2, male 73.5%, DM 17%, HTN 14.5%, CHD 2%, asthma 4.5% | Ivermectin 400 µgm/kg twice (second dose after one week) + SOC | SOC | Personal protective measures (PPM): hand hygiene, social distance measures, avoiding touching the eyes, nose, and, face masks, gloves, respiratory etiquette and self-isolation | NR |
| Krolewiecki et al;[152] 2020 | Peer-reviewed | Patients with mild to moderate COVID-19 (hospitalized) | 30 | 15 | Mean age 40.9 ± 12.5, male 56%, HTN 13.3%, DM 15.5%, COPD and asthma 11.1%, obesity 38% | Ivermectin 0.6 mg/kg for 5 days + SOC | SOC | NR | NR |

| | | | | | | | | | |
|---|---|---|---|---|---|---|---|---|---|
| Niaee et al;[153] 2020 | Pre-print | Patients with mild to severe COVID-19 (hospitalized) | 120 | 60 | Median age 56 ± 20, male 50% | Ivermectin 200 microg/kg once + SOC or Ivermectin 200 microg/kg daily for 3 days + SOC or Ivermectin 400 microg/kg once + SOC or Ivermectin 400 microg/kg once + 200 microg/kg daily for 2 days + SOC | SOC or placebo + SOC | Hydroxychloroquine 200mg/kg twice per day | NR |
| Ahmed et al;[154] 2020 | Peer-reviewed | Patients with mild to moderate COVID-19 (hospitalized patients) | 24 | 24 | Mean age 42, male 46%, | Ivermectin 12 mg a day for 5 days or ivermectin 12 mg a day for 5 days + doxycycline 100 mg and | placebo | NR | NR |
| Chaccour et al; 2020 | Peer-reviewed | Patients with mild COVID-19 (early within 3 days of onset, ambulatory) | 12 | 12 | Median age 26, male 50%, | Ivermectin 400 microg/kg | placebo | NR | NR |
| Chachar et al;[156] 2020 | Peer-reviewed | Patients with mild COVID-19 (ambulatory) | 25 | 25 | Mean age 41.8±15.7, male 62%, HTN 26%, DM 40%, obesity 12%, CHD 8% | Ivermectin 36mg once + SOC | SOC | NR | NR |
| Babalola et al;[157] 2020 | Peer-reviewed | Patients with mild to | 21 | 20 | Mean age 44.1 ± 14.7, male 69.4%, HTN 14.5%, | Ivermectin 12 mg a week + SOC, or | lopinavir-ritonavir + SOC | Zinc 62% | Steroids 3.2%, |

| | | | | | | | | | |
|---|---|---|---|---|---|---|---|---|---|
| | | severe COVID-19 (ambulatory and hospitalized) | | | DM 3.2% | ivermectin 24 mg a week for 2 weeks + SOC | | | |
| Kirti et al; 2020 | Pre-print | Patients with mild to moderate COVID-19 (hospitalized) | 55 | 57 | Mean age 52.5 ± 14.7, male 72.3%, HTN 34.8%, DM 35.7%, COPD 0.9%, asthma 0.9%, CHD 8.9%, CKD 2.7%, cerebrovascular disease 0%, cancer 5.4% | Ivermectin 24mg divided in two doses + SOC | Placebo + SOC | Steroids 100%, HCQ 100% | remdesivir 20.5%, tocilizumab 6.3%, convalescent plasma 13.4% |
| Chahla et al; 2020 | Pre-print | People exposed to SARS-COV-2 infection (ambulatory) | 117 | 117 | Median age 38 ± 12.5, male 42.7%, HTN 9%, DM 7.3%, CKD 2.1%, obesity 11.9% | Ivermectin + iota-carrageenan 12mg a week + 6 sprays a day for 4 weeks + SOC and | SOC | Standard biosecurity care and personal protective equipment (PPE) | NR |
| Mohan et al;[159] 2020 | Pre-print | Patients with mild to moderate COVID-19 (hospitalized) | 100 | 52 | Mean age 35.3 ± 10.4, male 88.8%, HTN 11.2%, DM 8.8%, CHD 0.8% | Ivermectin 12 mg (equivalent to 200 µg/kg) elixir + SOC or Ivermectin 24 mg (equivalent to 400 µg/kg) + SOC | Placebo + SOC | NR | Steroids 14.4%, remdesivir 1.6%, hydroxychloroquine 4%, azithromycin 11.2% |
| Samaha et al;[159] 2020 | Peer-reviewed | Patients with asymptomatic SARS-COV-2 infection (ambulatory) | 50 | 50 | Mean age: 31.78 ± 7.85, male 50%, HTN 8%, DM 6% | Ivermectin 0.2 mg/kg once + SOC | SOC | Zinc (30–50 mg/day) and Vitamin C (500 mg BID, twice daily) supplements | NR |

| | | | | | | | | | |
|---|---|---|---|---|---|---|---|---|---|
| Bukhari et al;[160] 2020 | Pre-print | Patients with mild to moderate COVID-19 (hospitalized) | 50 | 50 | Mean age: 40.6, males 73%, HTN 14%, DM 11.6%, CHD 5.8% | Ivermectin 12 mg once + SOC | SOC | vitamin C 500 mg once daily, vitamin D3 200,000 IU once weekly, Paracetamol 500 mg if necessary. | NR |
| Okumus et al;[161] 2021 | Peer-reviewed | Patients with severe COVID-19 (hospitalized) | 36 | 30 | Mean age 62 ± 12, male 60.6%, HTN 45%, DM 31.6%, COPD 15%, CHD 21.6%, cancer 1.6% | Ivermectin 0.2 mg/kg for 5 day + SOC | SOC | Hydroxychloroquine (2x400mg loading dose followed by 2x200mg for 5 days), favipiravir (2x1600mg loading dose followed by 2x600mg maintenance dose, po, total 5 days | NR |
| Beltran-Gonzalez et al;[126] 2021 | Pre-print | Patients with moderate to severe COVID-19 (hospitalized) | 36 | 37 | Mean age 54 ± 23.5, male 46.8%, HTN 19.1%, DM 9.6%, COPD 1%, CHD 7.4%, cerebrovascular disease 5.3% | Ivermectin 12-18 mg once | Placebo | dexamethasone, 6 mg IV every 24 hours, unfractioned heparin | Steroids 9.6%, lopinavir-ritonavir 44.7% |
| Lopez-Medina et al;[162] 2021 | Peer-reviewed | Patients with mild COVID-19 (ambulatory) | 238 | 238 | Median age 37 ± 19, male 42%, HTN 13.4%, DM 5.5%, COPD 3%, CHD 1.7%, cancer %, obesity 18.9% | Ivermectin 300 µg/kg a day for 5 days | Placebo | NR | Steroids 4.5% |
| Bermejo | Peer- | Patients with | 53 | 54 | Mean age 53.4 ± 15.6, | Ivermectin 42mg | CQ 900 mg for 1 day, | Azithromycin (500 | Steroids 98% |

| | | | | | | | | | |
|---|---|---|---|---|---|---|---|---|---|
| Galan et al;[128] 2021 | reviewed | severe to critical COVID-19 (hospitalized) | | | male 58.2%, HTN 43.4%, DM 28.1%, COPD 5.3%, CKD 2.5%, cancer 3%, obesity 37.5% | | and 450 mg for 4 days or HCQ 800 mg for 1 day and 400 mg for 4 days | mg 1× for 5 days) and ceftriaxone (1 g 2× for 7 days). Oseltamivir (75 mg 2× for 5 days) Proportions NR Steroids 98% | |
| Pott-Junior et al;[163] 2021 | Peer-reviewed | Patients with mild to severe COVID-19 (hospitalized) | 28 | 7 | Mean age 49.4 ± 14.6, male 45.2% | Ivermectin 100 + SOC or ivermectin 200 mc/kg + SOC or ivermectin 400 mcg/kg + SOC | SOC | NR. | Steroids 32.3% |
| Kishoria et al;[164] 2021 | Peer-reviewed | Patients with asymptomatic to mild COVID-19 (ambulatory) | 19 | 16 | Mean age 38 ±, male 66% | Ivermectin 12mg single dose + SOC | SOC | Hydroxychloroquine 400 mg twice a day, Paracetamol 500mg as required, Vitamin C 1 tab twice a day; for five days | NR |
| Seet et al;[129]; 2021 | Peer-reviewed | People exposed to SARS-COV-2 infection. (ambulatory) | 709 | 633 | Mean age 33, male 100%, hypertension 0.8%, diabetes 0.3% | Ivermectin 12mg once | vitamin C | NR | NR |
| Shahbaznejad et al; 2020 | Peer-reviewed | Patients with moderate to critical COVID-19 | 35 | 38 | Mean age 46.4 ± 22.5, male 52.2% | Ivermectin 0.2 mg/kg once + SOC | SOC | Chloroquine 75.4%, lopinavir-ritonavir 79.7%, antibiotics 89.8% | NR |

| | | | | | | | | | |
|---|---|---|---|---|---|---|---|---|---|
| | | (hospitalized) | | | | | | | |
| Abd-Elsalam S et al.; 2021 | Peer-reviewed | Patients with mild to moderate COVID-19 infection (hospitalized) | 82 | 82 | Mean age 40.9 ± 16.5, male 50%, HTN 20%, DM 16% | Ivermectin 12 mg a day for 3 days + SOC | SOC | Paracetamol, oxygen, fluids (according to the condition of the patient), empiric antibiotic,oseltamivir if needed (75 mg/12 h for 5 days) | NR |
| Biber et al; 2021 | Pre-print | Patients with mild recent onset COVID19 (ambulatory) | 47 | 42 | Mean age 35 ± 19, male 78.4% | Ivermectin 48 to 55 mg administered for three days + SOC | SOC | NR | NR |
| Faisal et al; 2021 | Peer-reviewed | Patients with mild COVID-19 (ambulatory) | 50 | 50 | Mean age 46 ± 3, male 80% | Ivermectin 12 mg a day for 5 days + SOC | SOC | Paracetamol (500mg if needed), Vit C (500mg once a day for 15 days), zinc (20mg twice a day for 15 days) and Vit D (injection PO 200,000units once) supplements. | NR |
| Vallejos et al; 2021 | Peer- | Patients with mild | 250 | 251 | Mean age 42.5 ± 15.5, male 52.7%, hypertension | Ivermectin 24 to 36mg (according to baseline | SOC | NR | NR |

| | | | | | | |
|---|---|---|---|---|---|---|
| | reviewed | COVID-19 (ambulatory) | | | 23.8%, diabetes 9.6%, COPD 2.8%, asthma 7.2%, CHD 1.8%, cancer 1.2% | bodyweight) divided in 2 doses |

SOC: standard of care; NR: not registered; HTN: arterial blood hypertension; DM: diabetes mellitus; CKD: chronic kidney disease; COPD: chronic obstructive pulmonary disease; CHD: coronary heart disease; CQ: chloroquine; HCQ: hydroxychloroquine.
