## Additional analysis plots for "Bias as a source of inconsistency in ivermectin trials for COVID-19: A systematic review"

**S9 Figure. Comparison: ivermectin vs. Standard of care; Outcome: mortality; Analysis: subgroups by intervention implemented.**

| Study | Experimental Events | Experimental Total | Control Events | Control Total | RR | 95%−CI | Weight (fixed) | Weight (random) |
|---|---|---|---|---|---|---|---|---|
| **Intervention = Ivermectin + Doxycycline** | | | | | | | | |
| Mahmud et al | 0 | 183 | 3 | 180 | 0.14 | [0.01; 2.70] | 3.8% | 3.1% |
| Hashim HA et al | 2 | 70 | 6 | 70 | 0.33 | [0.07; 1.60] | 6.5% | 7.8% |
| Fixed effect model | | 253 | | 250 | 0.26 | [0.07; 1.03] | 10.3% | −− |
| Random effects model | | | | | 0.28 | [0.07; 1.10] | −− | 10.9% |
| Heterogeneity: $I^2$ = 0%, $t^2$ = 0, $p$ = 0.61 | | | | | | | | |
| **Intervention = Ivermectin** | | | | | | | | |
| Elgazzar et al (mild) | 0 | 100 | 4 | 100 | 0.11 | [0.01; 2.04] | 4.9% | 3.2% |
| Elgazzar et al (severe) | 2 | 100 | 20 | 100 | 0.10 | [0.02; 0.42] | 21.7% | 8.7% |
| Niaee et al | 4 | 120 | 11 | 60 | 0.18 | [0.06; 0.55] | 15.9% | 11.2% |
| Kirti et al | 0 | 55 | 4 | 57 | 0.12 | [0.01; 2.09] | 4.8% | 3.2% |
| Shahbaznejad et al | 1 | 35 | 0 | 34 | 2.92 | [0.12; 69.14] | 0.5% | 2.8% |
| Okumus et al | 6 | 30 | 9 | 30 | 0.67 | [0.27; 1.64] | 9.7% | 13.0% |
| Beltran et al | 5 | 36 | 8 | 70 | 1.22 | [0.43; 3.45] | 5.9% | 11.7% |
| Lopez−Medina et al | 0 | 200 | 1 | 198 | 0.33 | [0.01; 8.05] | 1.6% | 2.7% |
| Bermejo Galan et al | 12 | 53 | 25 | 115 | 1.04 | [0.57; 1.91] | 17.1% | 15.8% |
| Abd−Elsalam et al | 3 | 82 | 4 | 82 | 0.75 | [0.17; 3.25] | 4.3% | 8.5% |
| Vallejos et al | 4 | 250 | 3 | 251 | 1.34 | [0.30; 5.92] | 3.2% | 8.3% |
| Fixed effect model | | 1061 | | 1097 | 0.53 | [0.38; 0.74] | 89.7% | −− |
| Random effects model | | | | | 0.53 | [0.29; 0.99] | −− | 89.1% |
| Heterogeneity: $I^2$ = 53%, $t^2$ = 0.4918, $p$ = 0.02 | | | | | | | | |
| **Fixed effect model** | | **1314** | | **1347** | **0.50** | **[0.36; 0.69]** | **100.0%** | **−−** |
| **Random effects model** | | | | | **0.50** | **[0.28; 0.88]** | **−−** | **100.0%** |

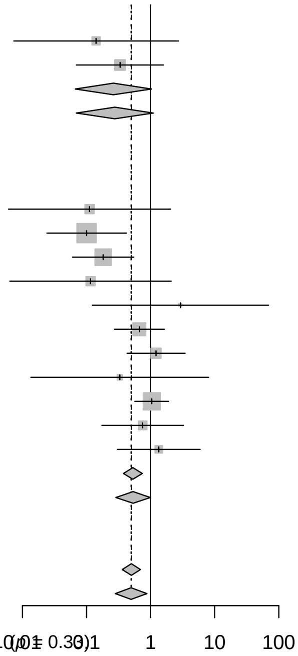


Heterogeneity: $I^2$ = 48%, $t^2$ = 0.4356, $p$ = 0.03
Test for subgroup differences (fixed effect): $c_1^2$ = 0.95, df = 1 ($p$ = 0.33)
Test for subgroup differences (random effects): $c_1^2$ = 0.72, df = 1 ($p$ = 0.40)

**S10 Figure. Comparison: ivermectin vs. Standard of care; Outcome: mortality; Analysis: subgroups by control implemented.**

| Study | Experimental Events | Experimental Total | Control Events | Control Total | RR | 95%−CI | Weight (fixed) | Weight (random) |
|---|---|---|---|---|---|---|---|---|
| **Control = SOC** | | | | | | | | |
| Mahmud et al | 0 | 183 | 3 | 180 | 0.14 | [0.01; 2.70] | 3.8% | 3.1% |
| Hashim HA et al | 2 | 70 | 6 | 70 | 0.33 | [0.07; 1.60] | 6.5% | 7.8% |
| Kirti et al | 0 | 55 | 4 | 57 | 0.12 | [0.01; 2.09] | 4.8% | 3.2% |
| Shahbaznejad et al | 1 | 35 | 0 | 34 | 2.92 | [0.12; 69.14] | 0.5% | 2.8% |
| Okumus et al | 6 | 30 | 9 | 30 | 0.67 | [0.27; 1.64] | 9.7% | 13.0% |
| Lopez−Medina et al | 0 | 200 | 1 | 198 | 0.33 | [0.01; 8.05] | 1.6% | 2.7% |
| Abd−Elsalam et al | 3 | 82 | 4 | 82 | 0.75 | [0.17; 3.25] | 4.3% | 8.5% |
| Vallejos et al | 4 | 250 | 3 | 251 | 1.34 | [0.30; 5.92] | 3.2% | 8.3% |
| Fixed effect model | | 905 | | 902 | 0.56 | [0.32; 0.98] | 34.6% | −− |
| Random effects model | | | | | 0.62 | [0.35; 1.10] | −− | 49.4% |
| Heterogeneity: $I^2$ = 0%, $t^2$ = 0, $p$ = 0.64 | | | | | | | | |
| **Control = HCQ** | | | | | | | | |
| Elgazzar et al (mild) | 0 | 100 | 4 | 100 | 0.11 | [0.01; 2.04] | 4.9% | 3.2% |
| Elgazzar et al (severe) | 2 | 100 | 20 | 100 | 0.10 | [0.02; 0.42] | 21.7% | 8.7% |
| Niaee et al | 4 | 120 | 11 | 60 | 0.18 | [0.06; 0.55] | 15.9% | 11.2% |
| Beltran et al | 5 | 36 | 8 | 70 | 1.22 | [0.43; 3.45] | 5.9% | 11.7% |
| Bermejo Galan et al | 12 | 53 | 25 | 115 | 1.04 | [0.57; 1.91] | 17.1% | 15.8% |
| Fixed effect model | | 409 | | 445 | 0.47 | [0.31; 0.70] | 65.4% | −− |
| Random effects model | | | | | 0.38 | [0.12; 1.14] | −− | 50.6% |
| Heterogeneity: $I^2$ = 78%, $t^2$ = 1.1313, $p$ < 0.01 | | | | | | | | |
| **Fixed effect model** | | **1314** | | **1347** | **0.50** | **[0.36; 0.69]** | **100.0%** | **−−** |
| **Random effects model** | | | | | **0.50** | **[0.28; 0.88]** | **−−** | **100.0%** |

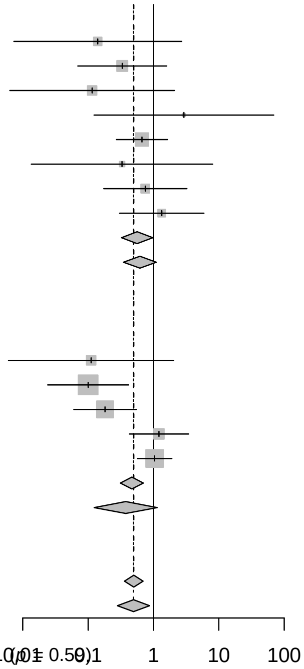


Heterogeneity: $I^2$ = 48%, $t^2$ = 0.4356, $p$ = 0.03
Test for subgroup differences (fixed effect): $c_1^2$ = 0.28, df = 1 ($p$ = 0.59)
Test for subgroup differences (random effects): $c_1^2$ = 0.61, df = 1 ($p$ = 0.43)

**S11 Figure. Comparison: ivermectin vs. Standard of care; Outcome: mortality; Analysis: subgroups by disease severity.**

| Study | Experimental Events | Experimental Total | Control Events | Control Total | RR | 95%−CI | Weight (fixed) | Weight (random) |
|---|---|---|---|---|---|---|---|---|
| **Severity = Mild to moderate** | | | | | | | | |
| Mahmud et al | 0 | 183 | 3 | 180 | 0.14 | [0.01; 2.70] | 3.8% | 3.1% |
| Elgazzar et al (mild) | 0 | 100 | 4 | 100 | 0.11 | [0.01; 2.04] | 4.9% | 3.2% |
| Kirti et al | 0 | 55 | 4 | 57 | 0.12 | [0.01; 2.09] | 4.8% | 3.2% |
| Lopez−Medina et al | 0 | 200 | 1 | 198 | 0.33 | [0.01; 8.05] | 1.6% | 2.7% |
| Abd−Elsalam et al | 3 | 82 | 4 | 82 | 0.75 | [0.17; 3.25] | 4.3% | 8.5% |
| Vallejos et al | 4 | 250 | 3 | 251 | 1.34 | [0.30; 5.92] | 3.2% | 8.3% |
| **Fixed effect model** | | 870 | | 868 | 0.43 | [0.20; 0.93] | 22.7% | −− |
| **Random effects model** | | | | | 0.54 | [0.23; 1.26] | −− | 29.0% |
| Heterogeneity: $I^2 = 0\%$, $\tau^2 = 0$, $p = 0.42$ | | | | | | | | |
| **Severity = Mixed** | | | | | | | | |
| Hashim HA et al | 2 | 70 | 6 | 70 | 0.33 | [0.07; 1.60] | 6.5% | 7.8% |
| Niaee et al | 4 | 120 | 11 | 60 | 0.18 | [0.06; 0.55] | 15.9% | 11.2% |
| Shahbaznejad et al | 1 | 35 | 0 | 34 | 2.92 | [0.12; 69.14] | 0.5% | 2.8% |
| Beltran et al | 5 | 36 | 8 | 70 | 1.22 | [0.43; 3.45] | 5.9% | 11.7% |
| **Fixed effect model** | | 261 | | 234 | 0.48 | [0.26; 0.88] | 28.8% | −− |
| **Random effects model** | | | | | 0.53 | [0.16; 1.68] | −− | 33.5% |
| Heterogeneity: $I^2 = 60\%$, $\tau^2 = 0.7813$, $p = 0.06$ | | | | | | | | |
| **Severity = Severe to critical** | | | | | | | | |
| Elgazzar et al (severe) | 2 | 100 | 20 | 100 | 0.10 | [0.02; 0.42] | 21.7% | 8.7% |
| Okumus et al | 6 | 30 | 9 | 30 | 0.67 | [0.27; 1.64] | 9.7% | 13.0% |
| Bermejo Galan et al | 12 | 53 | 25 | 115 | 1.04 | [0.57; 1.91] | 17.1% | 15.8% |
| **Fixed effect model** | | 183 | | 245 | 0.55 | [0.35; 0.85] | 48.5% | −− |
| **Random effects model** | | | | | 0.48 | [0.14; 1.58] | −− | 37.5% |
| Heterogeneity: $I^2 = 80\%$, $\tau^2 = 0.8726$, $p < 0.01$ | | | | | | | | |
| **Fixed effect model** | | **1314** | | **1347** | **0.50** | **[0.36; 0.69]** | **100.0%** | **−−** |
| **Random effects model** | | | | | **0.50** | **[0.28; 0.88]** | **−−** | **100.0%** |

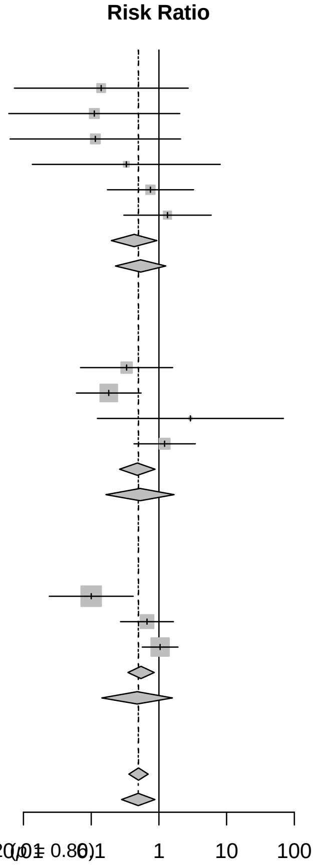


Heterogeneity: $I^2 = 48\%$, $\tau^2 = 0.4356$, $p = 0.03$

Test for subgroup differences (fixed effect): $\chi^2_2 = 0.31$, df = 2 ($p = 0.86$)

Test for subgroup differences (random effects): $\chi^2_2 = 0.02$, df = 2 ($p = 0.99$)

**S12 Figure. Comparison: ivermectin vs. Standard of care; Outcome: mechanical ventilation; Analysis: subgroups by intervention implemented.**

| Study | Experimental Events | Experimental Total | Control Events | Control Total | RR | 95%−CI | Weight (fixed) | Weight (random) |
|---|---|---|---|---|---|---|---|---|
| **Intervention = Ivermectin** | | | | | | | | |
| Krolewiecki et al | 1 | 20 | 0 | 12 | 1.83 | [0.08; 41.52] | 2.2% | 2.5% |
| Kirti et al | 1 | 55 | 5 | 57 | 0.21 | [0.03; 1.72] | 17.7% | 5.5% |
| Shahbaznejad et al | 2 | 35 | 1 | 34 | 1.94 | [0.18; 20.45] | 3.7% | 4.5% |
| Bermejo Galan et al | 12 | 53 | 24 | 115 | 1.08 | [0.59; 2.00] | 54.7% | 66.2% |
| Abd−Elsalam et al | 3 | 82 | 3 | 82 | 1.00 | [0.21; 4.81] | 10.8% | 10.0% |
| Vallejos et al | 4 | 250 | 3 | 251 | 1.34 | [0.30; 5.92] | 10.8% | 11.2% |
| **Fixed effect model** | | 495 | | 551 | 1.00 | [0.61; 1.62] | 100.0% | −− |
| **Random effects model** | | | | | 1.05 | [0.64; 1.72] | −− | 100.0% |
| Heterogeneity: $I^2 = 0\%$, $\tau^2 = 0$, $p = 0.73$ | | | | | | | | |
| **Fixed effect model** | | **495** | | **551** | **1.00** | **[0.61; 1.62]** | **100.0%** | **−−** |
| **Random effects model** | | | | | **1.05** | **[0.64; 1.72]** | **−−** | **100.0%** |

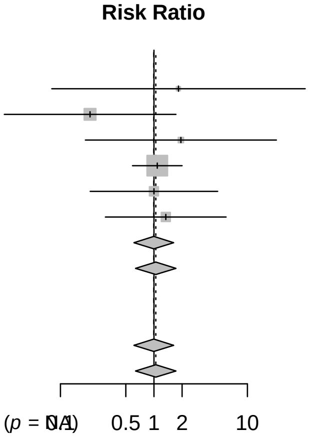


Heterogeneity: $I^2 = 0\%$, $\tau^2 = 0$, $p = 0.73$

Test for subgroup differences (fixed effect): $\chi^2_0 = 0.00$, df = 0 ($p$ = NA)

Test for subgroup differences (random effects): $\chi^2_0 = 0.00$, df = 0 ($p$ = NA)

**S13 Figure Comparison: ivermectin vs. Standard of care; Outcome: mechanical ventilation; Analysis: subgroups by control implemented.**

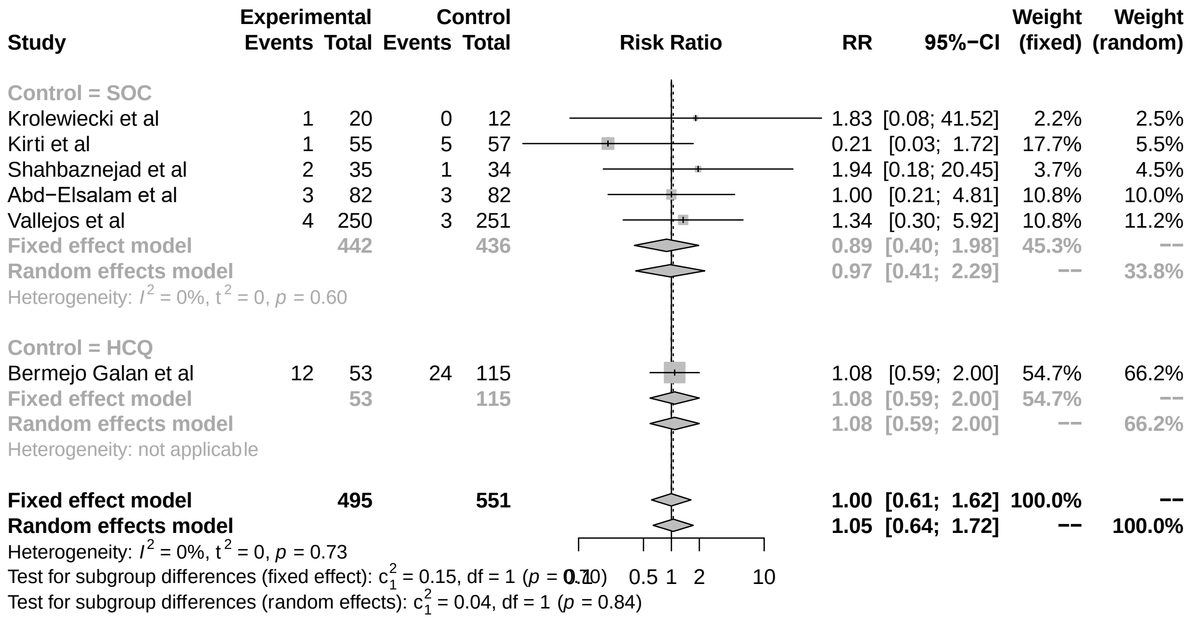


**S14 Figure. Comparison: ivermectin vs. Standard of care; Outcome: mechanical ventilation; Analysis: subgroups by disease severity.**

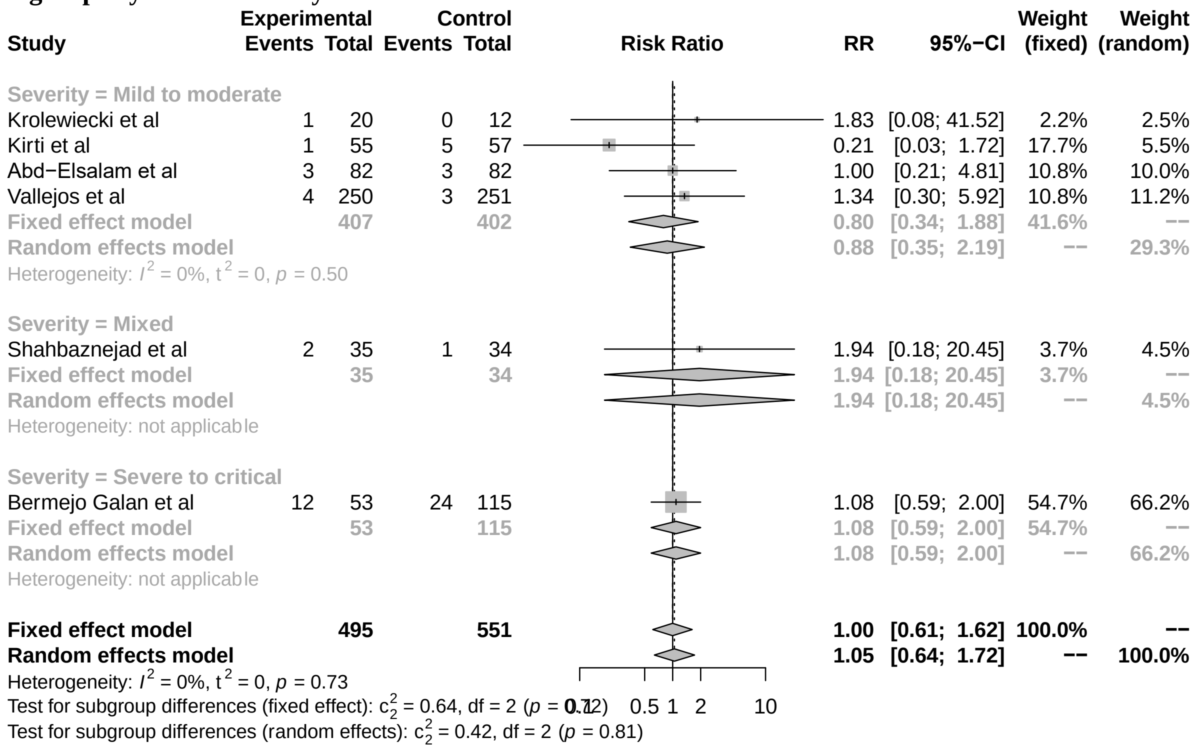


**S15 Figure. Comparison: ivermectin vs. Standard of care; Outcome: symptom resolution or improvement; Analysis: subgroups by intervention implemented.**

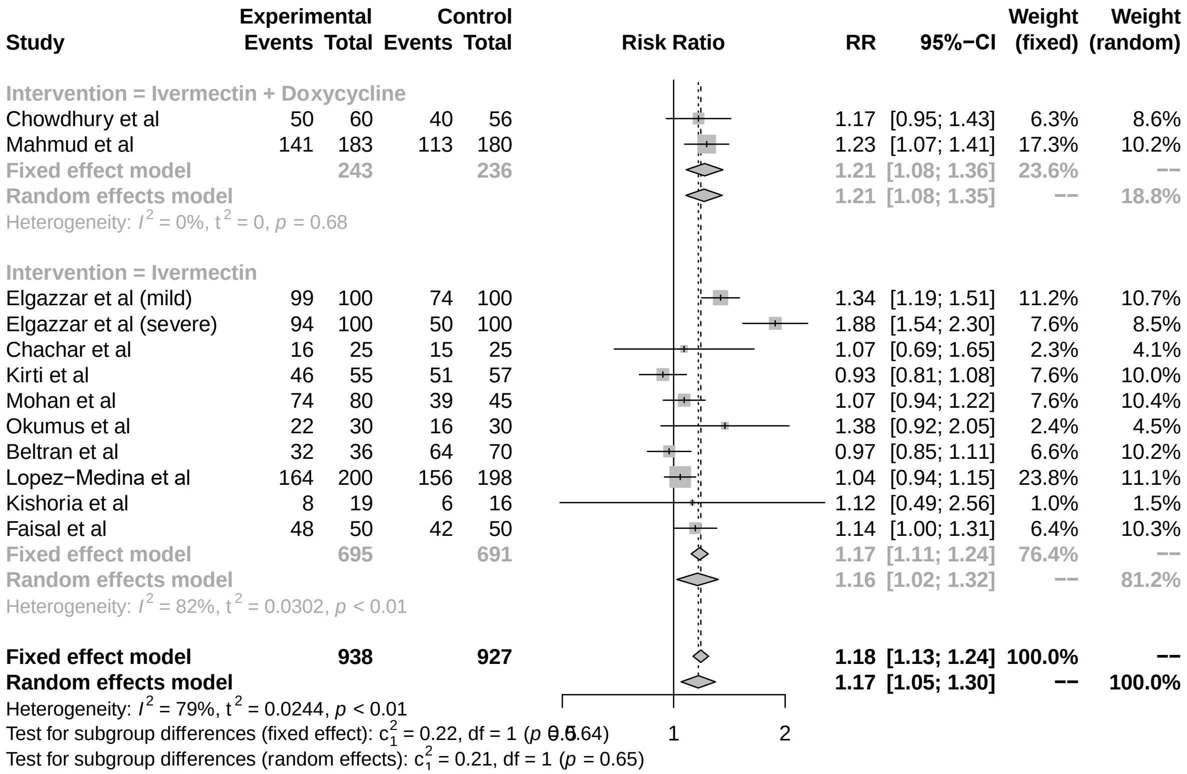


**S16 Figure. Comparison: ivermectin vs. Standard of care; Outcome: symptom resolution or improvement; Analysis: subgroups by control implemented.**

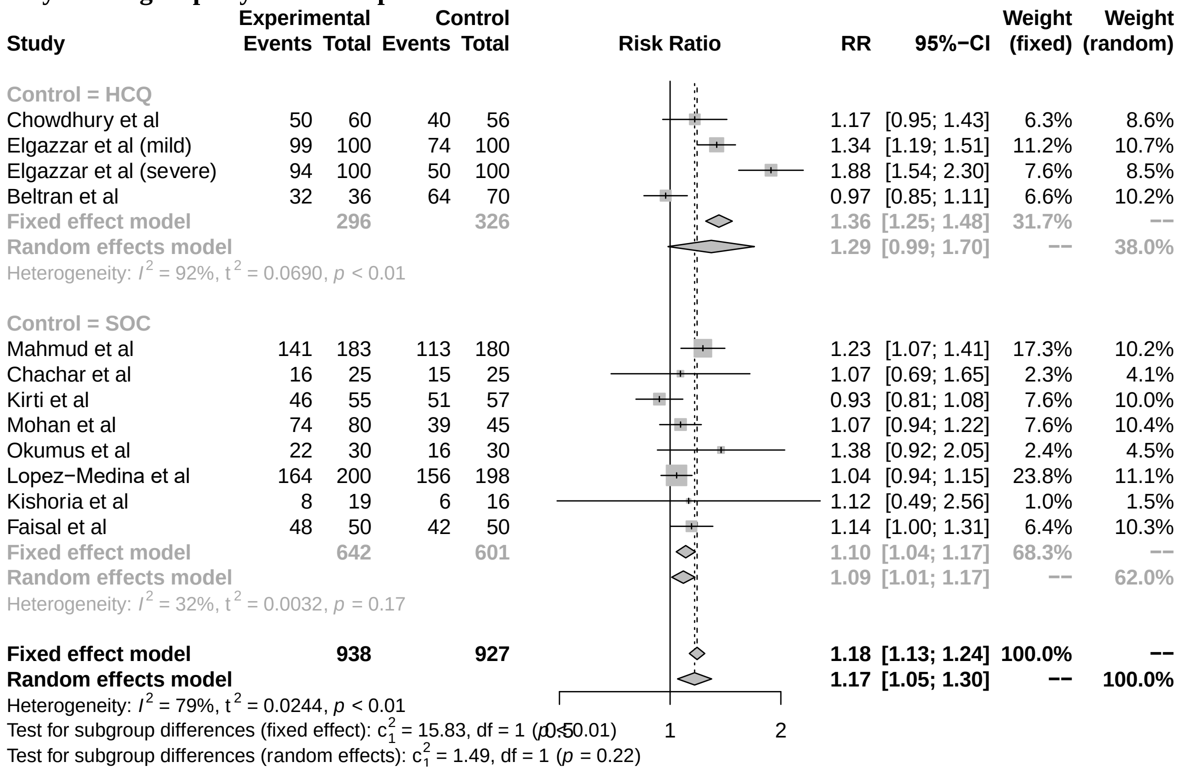


**S17 Figure. Comparison: ivermectin vs. Standard of care; Outcome: symptom resolution or improvement; Analysis: subgroups by disease severity.**

| Study | Experimental Events | Experimental Total | Control Events | Control Total | Risk Ratio | RR | 95%−CI | Weight (fixed) | Weight (random) |
|---|---|---|---|---|---|---|---|---|---|
| **Severity = Mild to moderate** | | | | | | | | | |
| Chowdhury et al | 50 | 60 | 40 | 56 | | 1.17 | [0.95; 1.43] | 6.3% | 8.6% |
| Mahmud et al | 141 | 183 | 113 | 180 | | 1.23 | [1.07; 1.41] | 17.3% | 10.2% |
| Elgazzar et al (mild) | 99 | 100 | 74 | 100 | | 1.34 | [1.19; 1.51] | 11.2% | 10.7% |
| Chachar et al | 16 | 25 | 15 | 25 | | 1.07 | [0.69; 1.65] | 2.3% | 4.1% |
| Kirti et al | 46 | 55 | 51 | 57 | | 0.93 | [0.81; 1.08] | 7.6% | 10.0% |
| Mohan et al | 74 | 80 | 39 | 45 | | 1.07 | [0.94; 1.22] | 7.6% | 10.4% |
| Lopez−Medina et al | 164 | 200 | 156 | 198 | | 1.04 | [0.94; 1.15] | 23.8% | 11.1% |
| Kishoria et al | 8 | 19 | 6 | 16 | | 1.12 | [0.49; 2.56] | 1.0% | 1.5% |
| Faisal et al | 48 | 50 | 42 | 50 | | 1.14 | [1.00; 1.31] | 6.4% | 10.3% |
| **Fixed effect model** | | **772** | | **727** | | **1.13** | **[1.07; 1.19]** | **83.4%** | **−−** |
| **Random effects model** | | | | | | **1.12** | **[1.03; 1.22]** | **−−** | **76.7%** |
| Heterogeneity: $I^2$ = 59%, $t^2$ = 0.0083, $p$ = 0.01 | | | | | | | | | |
| **Severity = Severe to critical** | | | | | | | | | |
| Elgazzar et al (severe) | 94 | 100 | 50 | 100 | | 1.88 | [1.54; 2.30] | 7.6% | 8.5% |
| Okumus et al | 22 | 30 | 16 | 30 | | 1.38 | [0.92; 2.05] | 2.4% | 4.5% |
| **Fixed effect model** | | **130** | | **130** | | **1.76** | **[1.47; 2.10]** | **10.0%** | **−−** |
| **Random effects model** | | | | | | **1.69** | **[1.26; 2.26]** | **−−** | **13.1%** |
| Heterogeneity: $I^2$ = 47%, $t^2$ = 0.0230, $p$ = 0.17 | | | | | | | | | |
| **Severity = Mixed** | | | | | | | | | |
| Beltran et al | 32 | 36 | 64 | 70 | | 0.97 | [0.85; 1.11] | 6.6% | 10.2% |
| **Fixed effect model** | | **36** | | **70** | | **0.97** | **[0.85; 1.11]** | **6.6%** | **−−** |
| **Random effects model** | | | | | | **0.97** | **[0.85; 1.11]** | **−−** | **10.2%** |
| Heterogeneity: not applicable | | | | | | | | | |
| **Fixed effect model** | | **938** | | **927** | | **1.18** | **[1.13; 1.24]** | **100.0%** | **−−** |
| **Random effects model** | | | | | | **1.17** | **[1.05; 1.30]** | **−−** | **100.0%** |

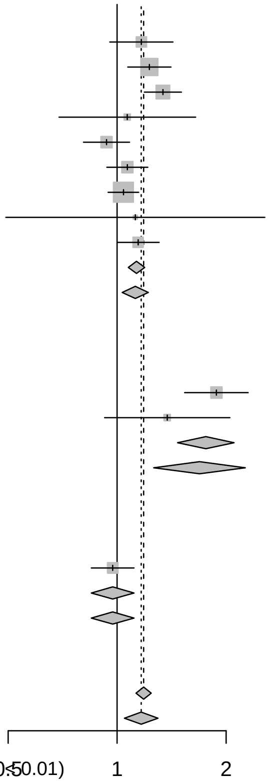


Heterogeneity: $I^2$ = 79%, $t^2$ = 0.0244, $p$ < 0.01
Test for subgroup differences (fixed effect): $c_2^2$ = 27.56, df = 2 ($p$ < 0.01)
Test for subgroup differences (random effects): $c_2^2$ = 11.68, df = 2 ($p$ < 0.01)

**S18 Figure. Comparison: ivermectin vs. Standard of care; Outcome: symptom resolution or improvement; Analysis: subgroups by outcome measurement time frame.**

| Study | Experimental Events | Experimental Total | Control Events | Control Total | Risk Ratio | RR | 95%−CI | Weight (fixed) | Weight (random) |
|---|---|---|---|---|---|---|---|---|---|
| **Follow: < 14 days** | | | | | | | | | |
| Chowdhury et al | 50 | 60 | 40 | 56 | | 1.17 | [0.95; 1.43] | 6.3% | 8.6% |
| Chachar et al | 16 | 25 | 15 | 25 | | 1.07 | [0.69; 1.65] | 2.3% | 4.1% |
| Kirti et al | 46 | 55 | 51 | 57 | | 0.93 | [0.81; 1.08] | 7.6% | 10.0% |
| Okumus et al | 22 | 30 | 16 | 30 | | 1.38 | [0.92; 2.05] | 2.4% | 4.5% |
| Beltran et al | 32 | 36 | 64 | 70 | | 0.97 | [0.85; 1.11] | 6.6% | 10.2% |
| Kishoria et al | 8 | 19 | 6 | 16 | | 1.12 | [0.49; 2.56] | 1.0% | 1.5% |
| Faisal et al | 48 | 50 | 42 | 50 | | 1.14 | [1.00; 1.31] | 6.4% | 10.3% |
| **Fixed effect model** | | **275** | | **304** | | **1.08** | **[0.99; 1.17]** | **32.5%** | **−−** |
| **Random effects model** | | | | | | **1.06** | **[0.96; 1.16]** | **−−** | **49.1%** |
| Heterogeneity: $I^2$ = 30%, $t^2$ = 0.0044, $p$ = 0.20 | | | | | | | | | |
| **Follow: > or = 14 days** | | | | | | | | | |
| Mahmud et al | 141 | 183 | 113 | 180 | | 1.23 | [1.07; 1.41] | 17.3% | 10.2% |
| Elgazzar et al (mild) | 99 | 100 | 74 | 100 | | 1.34 | [1.19; 1.51] | 11.2% | 10.7% |
| Elgazzar et al (severe) | 94 | 100 | 50 | 100 | | 1.88 | [1.54; 2.30] | 7.6% | 8.5% |
| Mohan et al | 74 | 80 | 39 | 45 | | 1.07 | [0.94; 1.22] | 7.6% | 10.4% |
| Lopez−Medina et al | 164 | 200 | 156 | 198 | | 1.04 | [0.94; 1.15] | 23.8% | 11.1% |
| **Fixed effect model** | | **663** | | **623** | | **1.24** | **[1.16; 1.31]** | **67.5%** | **−−** |
| **Random effects model** | | | | | | **1.26** | **[1.06; 1.50]** | **−−** | **50.9%** |
| Heterogeneity: $I^2$ = 89%, $t^2$ = 0.0338, $p$ < 0.01 | | | | | | | | | |
| **Fixed effect model** | | **938** | | **927** | | **1.18** | **[1.13; 1.24]** | **100.0%** | **−−** |
| **Random effects model** | | | | | | **1.17** | **[1.05; 1.30]** | **−−** | **100.0%** |

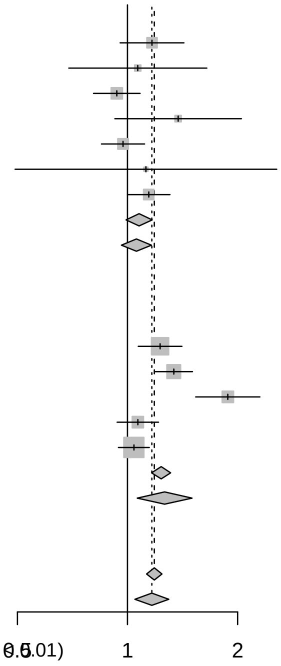


Heterogeneity: $I^2$ = 79%, $t^2$ = 0.0244, $p$ < 0.01
Test for subgroup differences (fixed effect): $c_1^2$ = 7.10, df = 1 ($p$ < 0.01)
Test for subgroup differences (random effects): $c_1^2$ = 3.13, df = 1 ($p$ = 0.08)

**S19 Figure. Comparison: ivermectin vs. Standard of care; Outcome: hospitalization; Analysis: subgroups by intervention implemented.**

| Study | Experimental Events | Experimental Total | Control Events | Control Total | RR | 95%−CI | Weight (fixed) | Weight (random) |
|---|---|---|---|---|---|---|---|---|
| Intervention = Ivermectin | | | | | | | | |
| Samaha et al | 0 | 50 | 3 | 50 | 0.14 | [0.01; 2.70] | 10.7% | 3.5% |
| Lopez−Medina et al | 4 | 200 | 6 | 198 | 0.66 | [0.19; 2.30] | 18.5% | 19.5% |
| Biber et al | 1 | 47 | 2 | 42 | 0.45 | [0.04; 4.75] | 6.5% | 5.5% |
| Vallejos et al | 14 | 250 | 21 | 251 | 0.67 | [0.35; 1.29] | 64.3% | 71.5% |
| Fixed effect model | | 547 | | 541 | 0.60 | [0.35; 1.03] | 100.0% | −− |
| Random effects model | | | | | 0.62 | [0.36; 1.07] | −− | 100.0% |
| Heterogeneity: $I^2 = 0\%$, $t^2 = 0$, $p = 0.77$ | | | | | | | | |
| **Fixed effect model** | | **547** | | **541** | **0.60** | **[0.35; 1.03]** | **100.0%** | **−−** |
| **Random effects model** | | | | | **0.62** | **[0.36; 1.07]** | **−−** | **100.0%** |

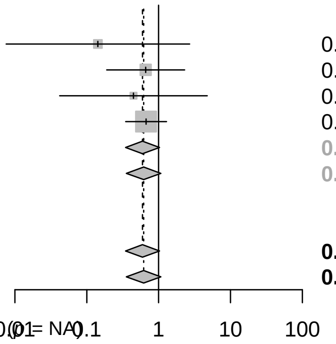


Heterogeneity: $I^2 = 0\%$, $t^2 = 0$, $p = 0.77$
Test for subgroup differences (fixed effect): $c_0^2 = 0.00$, df = 0 ($p$ = NA)
Test for subgroup differences (random effects): $c_0^2 = 0.00$, df = 0 ($p$ = NA)

**S20 Figure. Comparison: ivermectin vs. Standard of care; Outcome: hospitalization; Analysis: subgroups by control implemented.**

| Study | Experimental Events | Experimental Total | Control Events | Control Total | RR | 95%−CI | Weight (fixed) | Weight (random) |
|---|---|---|---|---|---|---|---|---|
| Control = SOC | | | | | | | | |
| Samaha et al | 0 | 50 | 3 | 50 | 0.14 | [0.01; 2.70] | 10.7% | 3.5% |
| Lopez−Medina et al | 4 | 200 | 6 | 198 | 0.66 | [0.19; 2.30] | 18.5% | 19.5% |
| Biber et al | 1 | 47 | 2 | 42 | 0.45 | [0.04; 4.75] | 6.5% | 5.5% |
| Vallejos et al | 14 | 250 | 21 | 251 | 0.67 | [0.35; 1.29] | 64.3% | 71.5% |
| Fixed effect model | | 547 | | 541 | 0.60 | [0.35; 1.03] | 100.0% | −− |
| Random effects model | | | | | 0.62 | [0.36; 1.07] | −− | 100.0% |
| Heterogeneity: $I^2 = 0\%$, $t^2 = 0$, $p = 0.77$ | | | | | | | | |
| **Fixed effect model** | | **547** | | **541** | **0.60** | **[0.35; 1.03]** | **100.0%** | **−−** |
| **Random effects model** | | | | | **0.62** | **[0.36; 1.07]** | **−−** | **100.0%** |

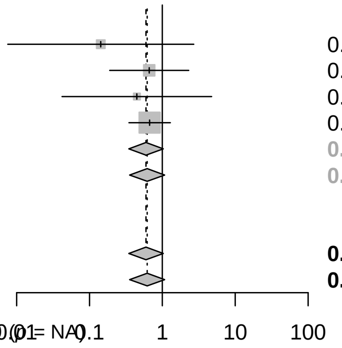


Heterogeneity: $I^2 = 0\%$, $t^2 = 0$, $p = 0.77$
Test for subgroup differences (fixed effect): $c_0^2 = 0.00$, df = 0 ($p$ = NA)
Test for subgroup differences (random effects): $c_0^2 = 0.00$, df = 0 ($p$ = NA)

**S21 Figure. Comparison: ivermectin vs. Standard of care; Outcome: hospitalization; Analysis: subgroups by disease severity.**

| Study | Experimental Events | Experimental Total | Control Events | Control Total | RR | 95%−CI | Weight (fixed) | Weight (random) |
|---|---|---|---|---|---|---|---|---|
| Severity = Mild to moderate | | | | | | | | |
| Samaha et al | 0 | 50 | 3 | 50 | 0.14 | [0.01; 2.70] | 10.7% | 3.5% |
| Lopez−Medina et al | 4 | 200 | 6 | 198 | 0.66 | [0.19; 2.30] | 18.5% | 19.5% |
| Biber et al | 1 | 47 | 2 | 42 | 0.45 | [0.04; 4.75] | 6.5% | 5.5% |
| Vallejos et al | 14 | 250 | 21 | 251 | 0.67 | [0.35; 1.29] | 64.3% | 71.5% |
| Fixed effect model | | 547 | | 541 | 0.60 | [0.35; 1.03] | 100.0% | −− |
| Random effects model | | | | | 0.62 | [0.36; 1.07] | −− | 100.0% |
| Heterogeneity: $I^2 = 0\%$, $t^2 = 0$, $p = 0.77$ | | | | | | | | |
| **Fixed effect model** | | **547** | | **541** | **0.60** | **[0.35; 1.03]** | **100.0%** | **−−** |
| **Random effects model** | | | | | **0.62** | **[0.36; 1.07]** | **−−** | **100.0%** |

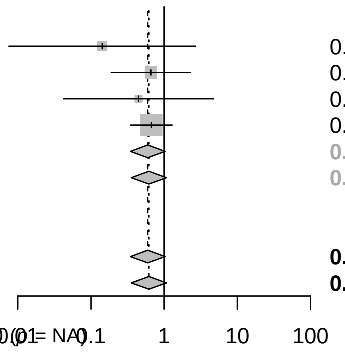


Heterogeneity: $I^2 = 0\%$, $t^2 = 0$, $p = 0.77$
Test for subgroup differences (fixed effect): $c_0^2 = 0.00$, df = 0 ($p$ = NA)
Test for subgroup differences (random effects): $c_0^2 = 0.00$, df = 0 ($p$ = NA)

**S22 Figure. Comparison: ivermectin vs. Standard of care; Outcome: infection; Analysis: subgroups by intervention implemented.**

| Study | Experimental Events | Experimental Total | Control Events | Control Total | RR | 95%−CI | Weight (fixed) | Weight (random) |
|---|---|---|---|---|---|---|---|---|
| Intervention = Ivermectin | | | | | | | | |
| Shouman et al | 15 | 203 | 59 | 101 | 0.13 | [0.08; 0.21] | 44.3% | 29.6% |
| Elgazzar et al (prophylaxis) | 2 | 100 | 10 | 100 | 0.20 | [0.04; 0.89] | 5.6% | 16.9% |
| Seet et al | 32 | 617 | 64 | 619 | 0.50 | [0.33; 0.76] | 36.0% | 30.8% |
| Fixed effect model | | 920 | | 820 | 0.29 | [0.21; 0.39] | 85.9% | −− |
| Random effects model | | | | | 0.24 | [0.08; 0.71] | −− | 77.3% |
| Heterogeneity: $I^2 = 88\%$, $\tau^2 = 0.7450$, $p < 0.01$ | | | | | | | | |
| Intervention = Ivermectin + iota−carrageenan | | | | | | | | |
| Chahla et al | 4 | 117 | 25 | 117 | 0.16 | [0.06; 0.45] | 14.1% | 22.7% |
| Fixed effect model | | 117 | | 117 | 0.16 | [0.06; 0.45] | 14.1% | −− |
| Random effects model | | | | | 0.16 | [0.06; 0.45] | −− | 22.7% |
| Heterogeneity: not applicable | | | | | | | | |
| **Fixed effect model** | | **1037** | | **937** | **0.27** | **[0.20; 0.36]** | **100.0%** | **−−** |
| **Random effects model** | | | | | **0.22** | **[0.09; 0.53]** | **−−** | **100.0%** |

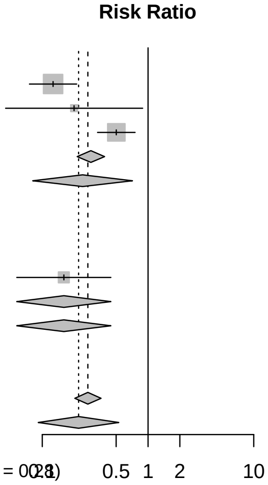


Heterogeneity: $I^2 = 84\%$, $\tau^2 = 0.6058$, $p < 0.01$
Test for subgroup differences (fixed effect): $\chi^2_1 = 1.17$, df = 1 ($p = 0.28$)
Test for subgroup differences (random effects): $\chi^2_1 = 0.29$, df = 1 ($p = 0.59$)

**S23 Figure. Comparison: ivermectin vs. Standard of care; Outcome: infection; Analysis: subgroups by control implemented.**

| Study | Experimental Events | Experimental Total | Control Events | Control Total | RR | 95%−CI | Weight (fixed) | Weight (random) |
|---|---|---|---|---|---|---|---|---|
| Control = SOC | | | | | | | | |
| Shouman et al | 15 | 203 | 59 | 101 | 0.13 | [0.08; 0.21] | 44.3% | 29.6% |
| Chahla et al | 4 | 117 | 25 | 117 | 0.16 | [0.06; 0.45] | 14.1% | 22.7% |
| Seet et al | 32 | 617 | 64 | 619 | 0.50 | [0.33; 0.76] | 36.0% | 30.8% |
| Fixed effect model | | 937 | | 837 | 0.27 | [0.21; 0.37] | 94.4% | −− |
| Random effects model | | | | | 0.22 | [0.08; 0.62] | −− | 83.1% |
| Heterogeneity: $I^2 = 89\%$, $\tau^2 = 0.7047$, $p < 0.01$ | | | | | | | | |
| Control = HCQ | | | | | | | | |
| Elgazzar et al (prophylaxis) | 2 | 100 | 10 | 100 | 0.20 | [0.04; 0.89] | 5.6% | 16.9% |
| Fixed effect model | | 100 | | 100 | 0.20 | [0.04; 0.89] | 5.6% | −− |
| Random effects model | | | | | 0.20 | [0.04; 0.89] | −− | 16.9% |
| Heterogeneity: not applicable | | | | | | | | |
| **Fixed effect model** | | **1037** | | **937** | **0.27** | **[0.20; 0.36]** | **100.0%** | **−−** |
| **Random effects model** | | | | | **0.22** | **[0.09; 0.53]** | **−−** | **100.0%** |

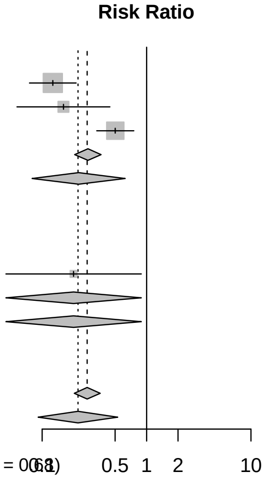


Heterogeneity: $I^2 = 84\%$, $\tau^2 = 0.6058$, $p < 0.01$
Test for subgroup differences (fixed effect): $\chi^2_1 = 0.17$, df = 1 ($p = 0.68$)
Test for subgroup differences (random effects): $\chi^2_1 = 0.01$, df = 1 ($p = 0.90$)

**S24 Figure. Comparison: ivermectin vs. Standard of care; Outcome: viral clearance; Analysis: subgroups by intervention implemented.**

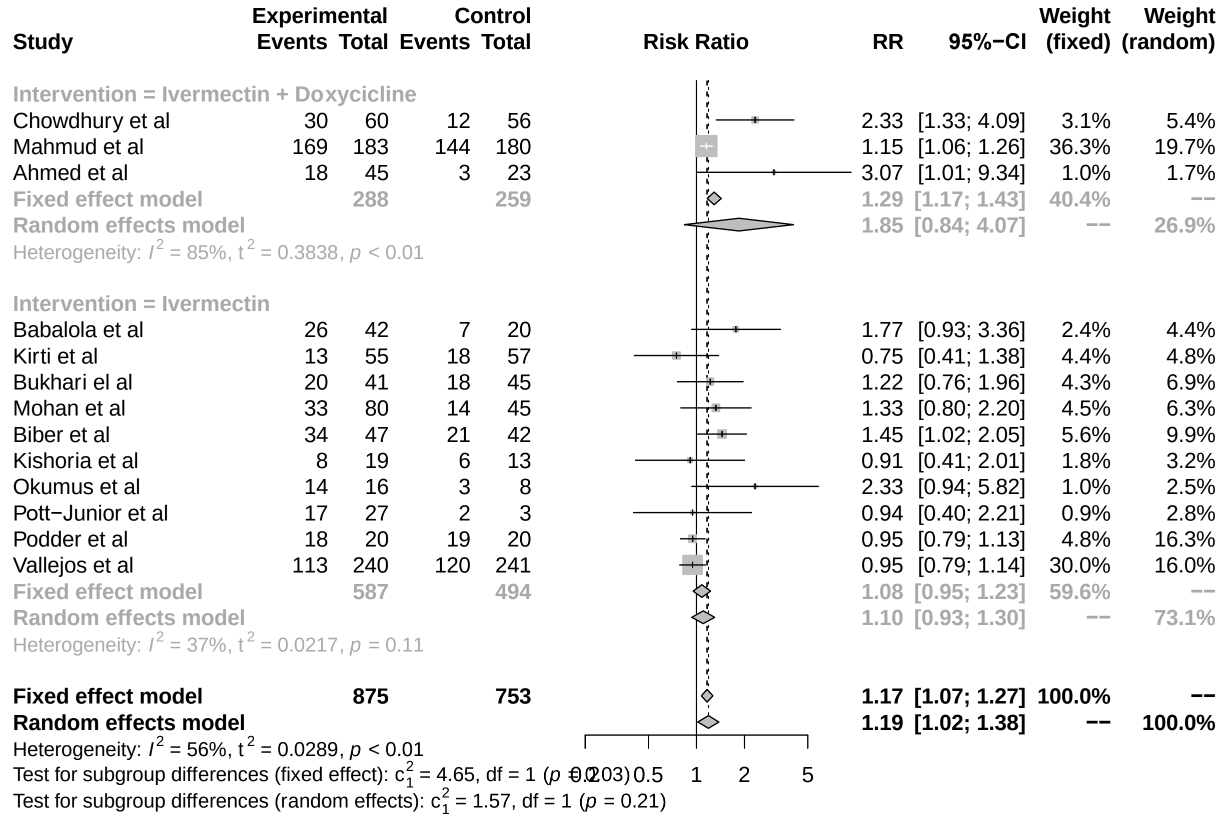


**S25 Figure. Comparison: ivermectin vs. Standard of care; Outcome: viral clearance; Analysis: subgroups by control implemented.**

| Study | Experimental Events | Experimental Total | Control Events | Control Total | RR | 95%−CI | Weight (fixed) | Weight (random) |
|---|---|---|---|---|---|---|---|---|
| **Control = HCQ** | | | | | | | | |
| Chowdhury et al | 30 | 60 | 12 | 56 | 2.33 | [1.33; 4.09] | 3.1% | 5.4% |
| **Fixed effect model** | | **60** | | **56** | **2.33** | **[1.33; 4.09]** | **3.1%** | **−−** |
| **Random effects model** | | | | | **2.33** | **[1.33; 4.09]** | **−−** | **5.4%** |
| Heterogeneity: not applicable | | | | | | | | |
| **Control = SOC** | | | | | | | | |
| Mahmud et al | 169 | 183 | 144 | 180 | 1.15 | [1.06; 1.26] | 36.3% | 19.7% |
| Ahmed et al | 18 | 45 | 3 | 23 | 3.07 | [1.01; 9.34] | 1.0% | 1.7% |
| Kirti et al | 13 | 55 | 18 | 57 | 0.75 | [0.41; 1.38] | 4.4% | 4.8% |
| Bukhari el al | 20 | 41 | 18 | 45 | 1.22 | [0.76; 1.96] | 4.3% | 6.9% |
| Mohan et al | 33 | 80 | 14 | 45 | 1.33 | [0.80; 2.20] | 4.5% | 6.3% |
| Biber et al | 34 | 47 | 21 | 42 | 1.45 | [1.02; 2.05] | 5.6% | 9.9% |
| Kishoria et al | 8 | 19 | 6 | 13 | 0.91 | [0.41; 2.01] | 1.8% | 3.2% |
| Okumus et al | 14 | 16 | 3 | 8 | 2.33 | [0.94; 5.82] | 1.0% | 2.5% |
| Pott−Junior et al | 17 | 27 | 2 | 3 | 0.94 | [0.40; 2.21] | 0.9% | 2.8% |
| Podder et al | 18 | 20 | 19 | 20 | 0.95 | [0.79; 1.13] | 4.8% | 16.3% |
| Vallejos et al | 113 | 240 | 120 | 241 | 0.95 | [0.79; 1.14] | 30.0% | 16.0% |
| **Fixed effect model** | | **773** | | **677** | **1.11** | **[1.02; 1.21]** | **94.5%** | **−−** |
| **Random effects model** | | | | | **1.10** | **[0.97; 1.26]** | **−−** | **90.1%** |
| Heterogeneity: $I^2 = 42\%$, $t^2 = 0.0148$, $p = 0.07$ | | | | | | | | |
| **Control = Lopinavir−Ritonavir** | | | | | | | | |
| Babalola et al | 26 | 42 | 7 | 20 | 1.77 | [0.93; 3.36] | 2.4% | 4.4% |
| **Fixed effect model** | | **42** | | **20** | **1.77** | **[0.93; 3.36]** | **2.4%** | **−−** |
| **Random effects model** | | | | | **1.77** | **[0.93; 3.36]** | **−−** | **4.4%** |
| Heterogeneity: not applicable | | | | | | | | |
| **Fixed effect model** | | **875** | | **753** | **1.17** | **[1.07; 1.27]** | **100.0%** | **−−** |
| **Random effects model** | | | | | **1.19** | **[1.02; 1.38]** | **−−** | **100.0%** |

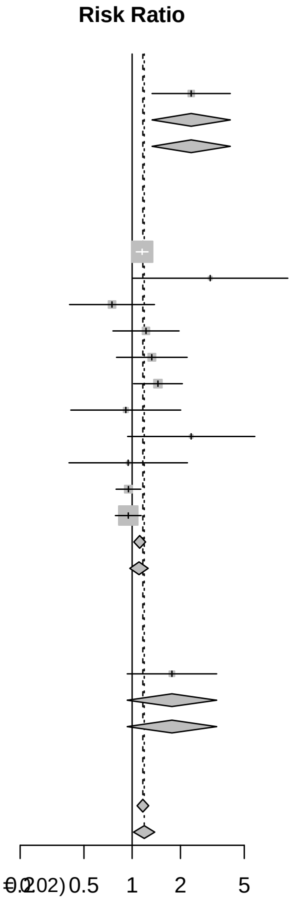


Heterogeneity: $I^2 = 56\%$, $t^2 = 0.0289$, $p < 0.01$
Test for subgroup differences (fixed effect): $c^2_2 = 8.35$, df = 2 ($p = 0.02$)
Test for subgroup differences (random effects): $c^2_2 = 8.16$, df = 2 ($p = 0.02$)

**S26 Figure. Comparison: ivermectin vs. Standard of care; Outcome: viral clearance; Analysis: subgroups by disease severity.**

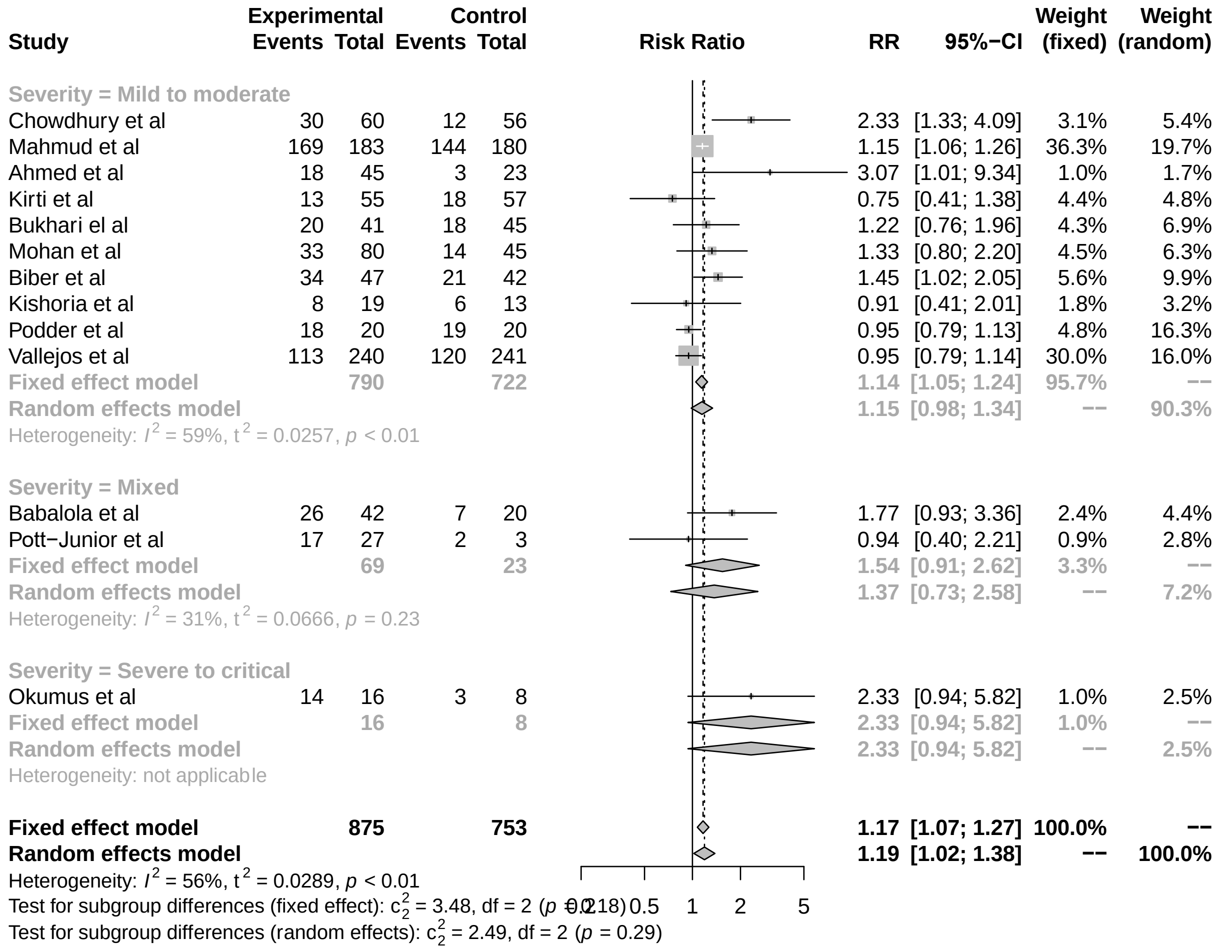


**S27 Figure. Comparison: ivermectin vs. Standard of care; Outcome: viral clearance; Analysis: subgroups by outcome measurement time frame.**

| Study | Experimental Events | Experimental Total | Control Events | Control Total | RR | 95%−CI | Weight (fixed) | Weight (random) |
|---|---|---|---|---|---|---|---|---|
| **Follow: > 7 days** | | | | | | | | |
| Chowdhury et al | 30 | 60 | 12 | 56 | 2.33 | [1.33; 4.09] | 3.1% | 5.4% |
| Mahmud et al | 169 | 183 | 144 | 180 | 1.15 | [1.06; 1.26] | 36.3% | 19.7% |
| Podder et al | 18 | 20 | 19 | 20 | 0.95 | [0.79; 1.13] | 4.8% | 16.3% |
| **Fixed effect model** | | **263** | | **256** | **1.21** | **[1.11; 1.33]** | **44.2%** | **−−** |
| **Random effects model** | | | | | **1.21** | **[0.89; 1.65]** | **−−** | **41.5%** |
| Heterogeneity: $I^2 = 86\%$, $\tau^2 = 0.0554$, $p < 0.01$ | | | | | | | | |
| **Follow: < or = 7 days** | | | | | | | | |
| Ahmed et al | 18 | 45 | 3 | 23 | 3.07 | [1.01; 9.34] | 1.0% | 1.7% |
| Babalola et al | 26 | 42 | 7 | 20 | 1.77 | [0.93; 3.36] | 2.4% | 4.4% |
| Kirti et al | 13 | 55 | 18 | 57 | 0.75 | [0.41; 1.38] | 4.4% | 4.8% |
| Bukhari el al | 20 | 41 | 18 | 45 | 1.22 | [0.76; 1.96] | 4.3% | 6.9% |
| Mohan et al | 33 | 80 | 14 | 45 | 1.33 | [0.80; 2.20] | 4.5% | 6.3% |
| Biber et al | 34 | 47 | 21 | 42 | 1.45 | [1.02; 2.05] | 5.6% | 9.9% |
| Kishoria et al | 8 | 19 | 6 | 13 | 0.91 | [0.41; 2.01] | 1.8% | 3.2% |
| Okumus et al | 14 | 16 | 3 | 8 | 2.33 | [0.94; 5.82] | 1.0% | 2.5% |
| Pott−Junior et al | 17 | 27 | 2 | 3 | 0.94 | [0.40; 2.21] | 0.9% | 2.8% |
| Vallejos et al | 113 | 240 | 120 | 241 | 0.95 | [0.79; 1.14] | 30.0% | 16.0% |
| **Fixed effect model** | | **612** | | **497** | **1.13** | **[0.99; 1.29]** | **55.8%** | **−−** |
| **Random effects model** | | | | | **1.21** | **[0.98; 1.51]** | **−−** | **58.5%** |
| Heterogeneity: $I^2 = 42\%$, $\tau^2 = 0.0444$, $p = 0.08$ | | | | | | | | |
| **Fixed effect model** | | **875** | | **753** | **1.17** | **[1.07; 1.27]** | **100.0%** | **−−** |
| **Random effects model** | | | | | **1.19** | **[1.02; 1.38]** | **−−** | **100.0%** |

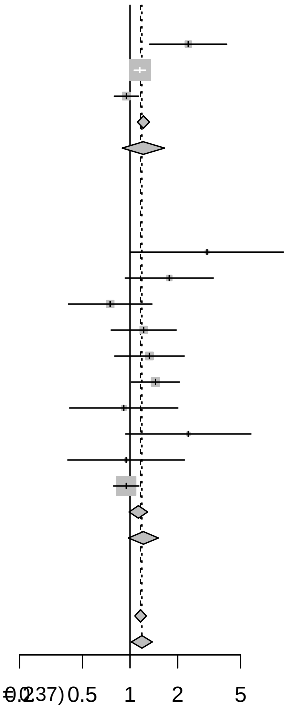


Heterogeneity: $I^2 = 56\%$, $\tau^2 = 0.0289$, $p < 0.01$
Test for subgroup differences (fixed effect): $c_1^2 = 0.81$, df = 1 ($p = 0.37$)
Test for subgroup differences (random effects): $c_1^2 = 0.00$, df = 1 ($p = 0.99$)

**S28 Figure. Comparison: ivermectin vs. Standard of care; Outcome: adverse events; Analysis: subgroups by intervention implemented.**

| Study | Experimental Events | Experimental Total | Control Events | Control Total | RR | 95%−CI | Weight (fixed) | Weight (random) |
|---|---|---|---|---|---|---|---|---|
| **Intervention = Ivermectin + Doxycycline** | | | | | | | | |
| Mahmud et al | 2 | 183 | 0 | 180 | 4.92 | [0.24; 101.73] | 10.3% | 15.1% |
| **Fixed effect model** | | **183** | | **180** | **4.92** | **[0.24; 101.74]** | **10.3%** | **−−** |
| **Random effects model** | | | | | **4.92** | **[0.24; 101.73]** | **−−** | **15.1%** |
| Heterogeneity: not applicable | | | | | | | | |
| **Intervention = Ivermectin** | | | | | | | | |
| Krolewiecki et al | 1 | 20 | 0 | 12 | 1.83 | [0.08; 41.52] | 12.7% | 14.2% |
| Lopez−Medina et al | 2 | 200 | 2 | 198 | 0.99 | [0.14; 6.96] | 41.2% | 36.4% |
| Pott−Junior et al | 3 | 27 | 1 | 4 | 0.44 | [0.06; 3.30] | 35.7% | 34.4% |
| **Fixed effect model** | | **247** | | **214** | **0.90** | **[0.25; 3.19]** | **89.7%** | **−−** |
| **Random effects model** | | | | | **0.79** | **[0.22; 2.84]** | **−−** | **84.9%** |
| Heterogeneity: $I^2 = 0\%$, $\tau^2 = 0$, $p = 0.71$ | | | | | | | | |
| **Fixed effect model** | | **430** | | **394** | **1.31** | **[0.43; 4.02]** | **100.0%** | **−−** |
| **Random effects model** | | | | | **1.04** | **[0.32; 3.38]** | **−−** | **100.0%** |

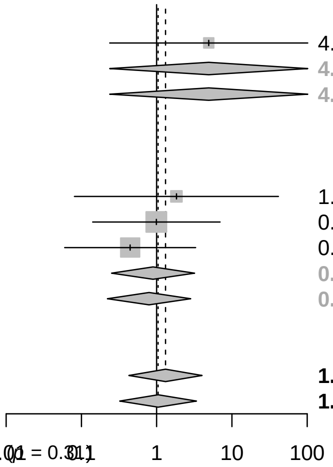


Heterogeneity: $I^2 = 0\%$, $\tau^2 = 0$, $p = 0.58$
Test for subgroup differences (fixed effect): $c_1^2 = 1.03$, df = 1 ($p = 0.31$)
Test for subgroup differences (random effects): $c_1^2 = 1.18$, df = 1 ($p = 0.28$)

**S29 Figure. Comparison: ivermectin vs. Standard of care; Outcome: adverse events; Analysis: subgroups by control implemented.**

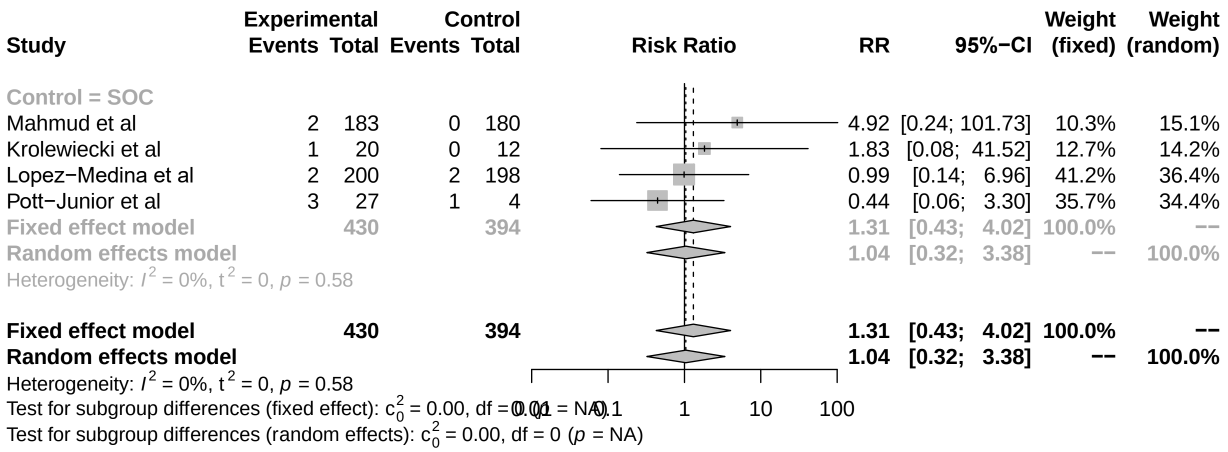


**S30 Figure. Comparison: ivermectin vs. Standard of care; Outcome: adverse events; Analysis: subgroups by disease severity.**

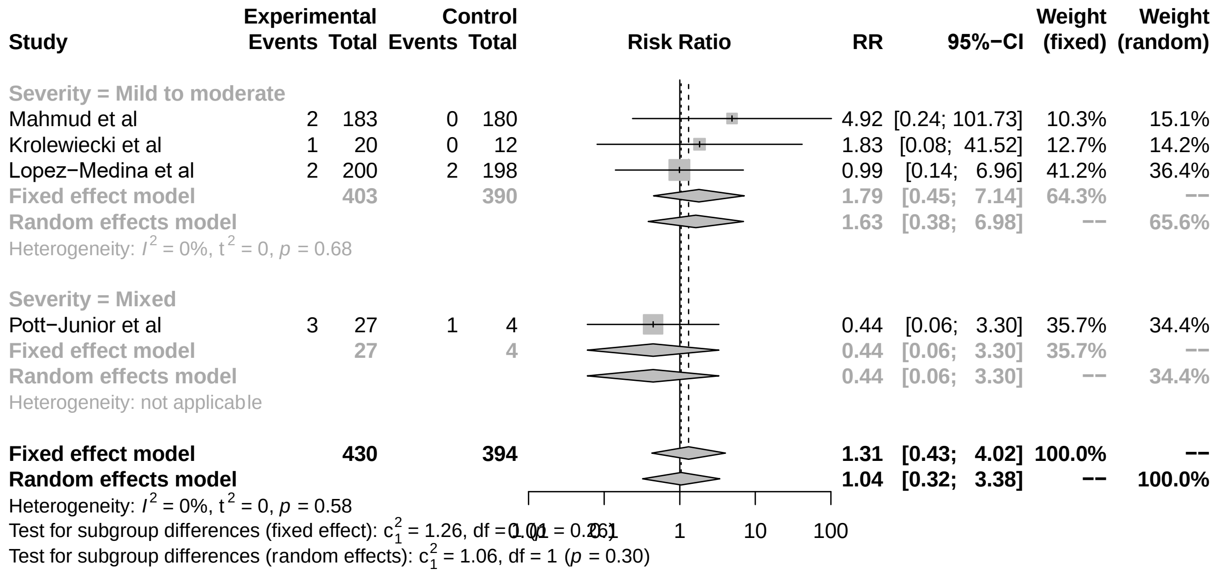


**S31 Figure. Comparison: ivermectin vs. Standard of care; Outcome: mortality; Analysis: risk of publication bias.**

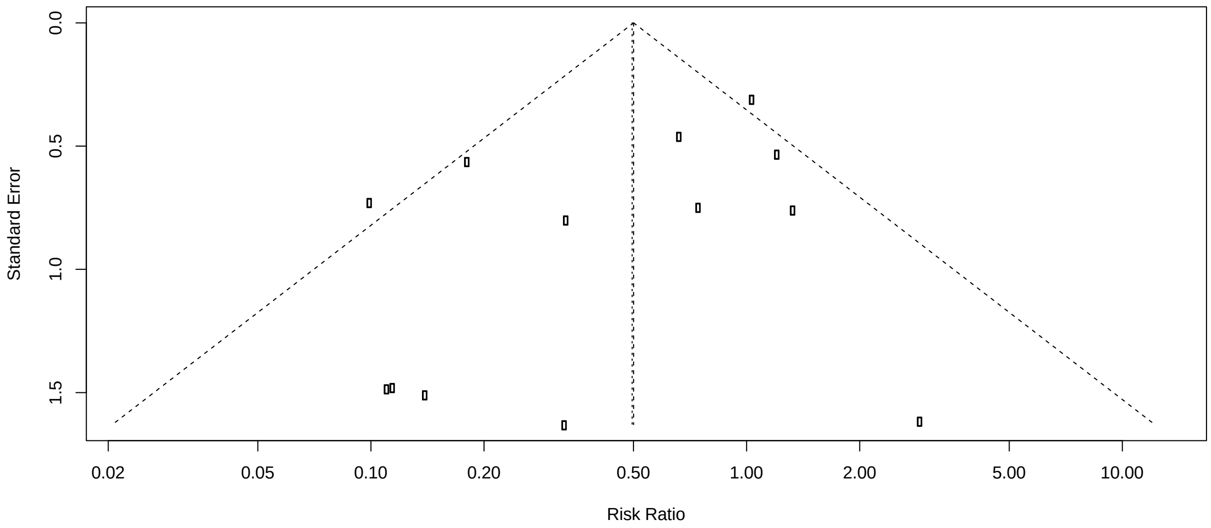


**S32 Figure. Comparison: ivermectin vs. Standard of care; Outcome: symptom resolution or improvement; Analysis: risk of publication bias.**

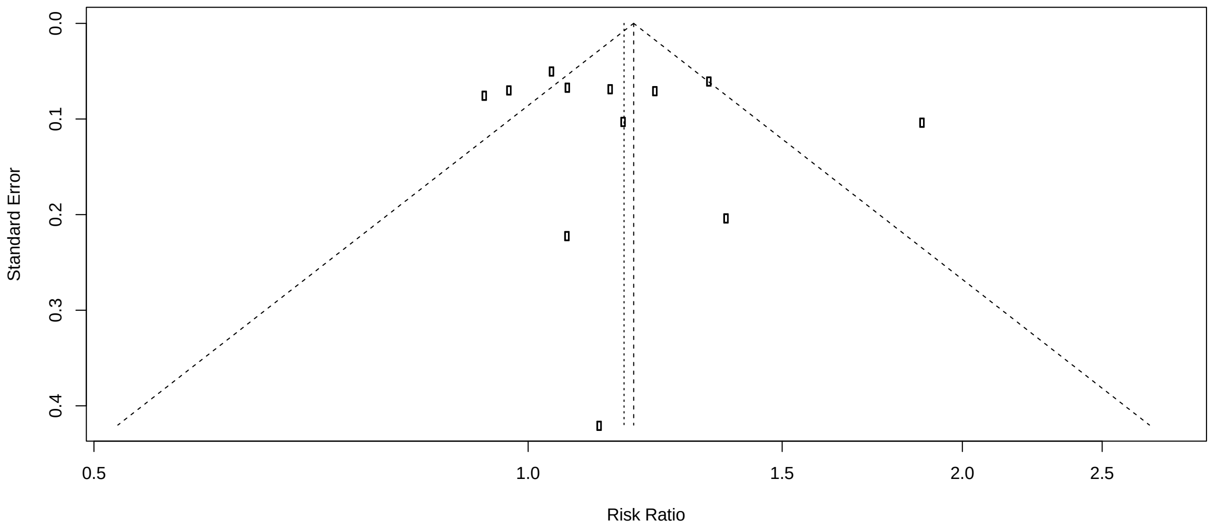


**S33 Figure. Comparison: ivermectin vs. Standard of care; Outcome: viral clearance; Analysis: risk of publication bias.**

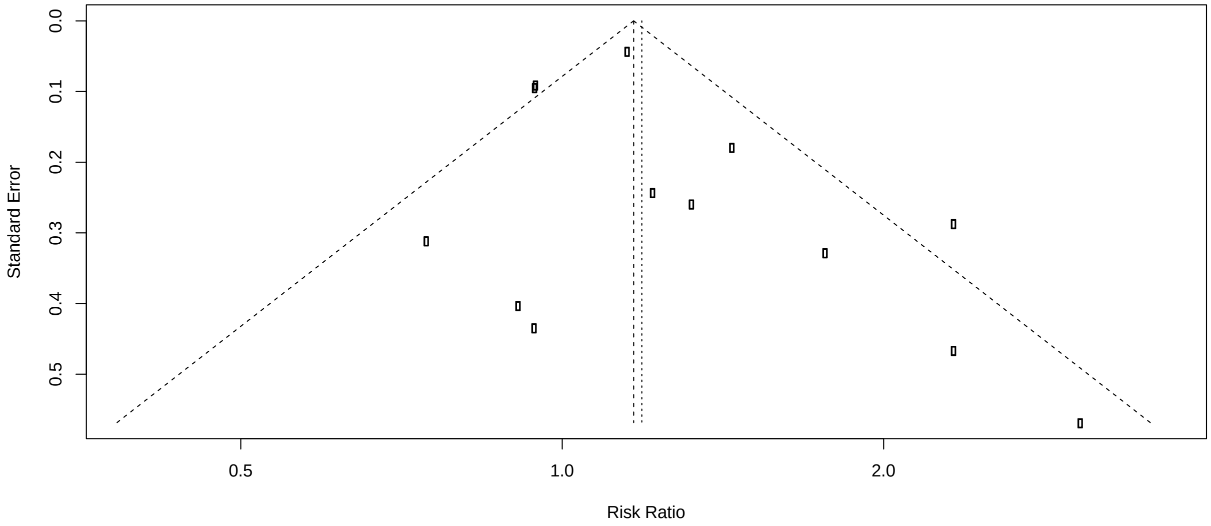
0.0
0.1
0.2
0.3
0.4
0.5
Standard Error
0.5
1.0
2.0
Risk Ratio
