## Supplementary material for "Bias as a source of inconsistency in ivermectin trials for COVID-19: A systematic review": Baseline prognostic factors balance between study arms

#### S2 Figure. Baseline prognostic factors balance between study arms

##### Male gender

| Study | Experimental Events | Experimental Total | Control Events | Control Total | RR | 95%−CI | Weight (fixed) | Weight (random) |
|---|---|---|---|---|---|---|---|---|
| Shouman et al | 106 | 203 | 50 | 101 | 1.05 | [0.83; 1.34] | 5.7% | 3.8% |
| Chowdhury et al | 43 | 60 | 47 | 56 | 0.85 | [0.70; 1.04] | 4.2% | 5.5% |
| Podder et al | 23 | 32 | 21 | 30 | 1.03 | [0.75; 1.41] | 1.9% | 2.1% |
| Mahmud et al | 123 | 200 | 112 | 200 | 1.10 | [0.93; 1.29] | 9.6% | 7.8% |
| Hashim et al | 37 | 70 | 36 | 70 | 1.03 | [0.75; 1.41] | 3.1% | 2.1% |
| Elgazzar et al − Mild | 72 | 100 | 67 | 100 | 1.07 | [0.89; 1.29] | 5.7% | 6.3% |
| Elgazzar et al − Severe | 68 | 100 | 74 | 100 | 0.92 | [0.77; 1.10] | 6.3% | 6.7% |
| Elgazzar et al − Prophylaxis | 75 | 100 | 72 | 100 | 1.04 | [0.88; 1.23] | 6.1% | 7.7% |
| Krolewiecki et al | 15 | 30 | 10 | 15 | 0.75 | [0.45; 1.24] | 1.1% | 0.8% |
| Niaee et al | 78 | 126 | 30 | 49 | 1.01 | [0.78; 1.31] | 3.7% | 3.1% |
| Chaccour et al | 7 | 12 | 5 | 12 | 1.40 | [0.61; 3.19] | 0.4% | 0.3% |
| Chachar et al | 17 | 25 | 14 | 25 | 1.21 | [0.78; 1.88] | 1.2% | 1.1% |
| Babalola et al | 29 | 42 | 14 | 20 | 0.99 | [0.69; 1.40] | 1.6% | 1.7% |
| Kirti et al | 40 | 55 | 41 | 57 | 1.01 | [0.80; 1.27] | 3.4% | 4.1% |
| Okumus et al | 21 | 30 | 19 | 30 | 1.11 | [0.77; 1.58] | 1.6% | 1.6% |
| Beltran et al | 21 | 36 | 23 | 36 | 0.91 | [0.63; 1.32] | 2.0% | 1.6% |
| Lopez−Medina et al | 78 | 200 | 89 | 198 | 0.87 | [0.69; 1.09] | 7.6% | 4.0% |
| Bermejo Galan et al | 31 | 53 | 64 | 115 | 1.05 | [0.79; 1.39] | 3.4% | 2.7% |
| Shahbaznejad et al | 18 | 35 | 18 | 34 | 0.97 | [0.62; 1.53] | 1.6% | 1.0% |
| Abd−Elsalam et al | 37 | 82 | 45 | 82 | 0.82 | [0.60; 1.12] | 3.8% | 2.2% |
| Biber et al | 36 | 47 | 33 | 42 | 0.97 | [0.78; 1.22] | 3.0% | 4.3% |
| Chahala et al | 52 | 117 | 48 | 117 | 1.08 | [0.80; 1.46] | 4.1% | 2.4% |
| Bukhari et al | 37 | 41 | 36 | 45 | 1.13 | [0.94; 1.35] | 2.9% | 6.8% |
| Pott−Junior | 14 | 27 | 0 | 7 | 7.91 | [0.53; 117.82] | 0.1% | 0.0% |
| Kishoria et al | 14 | 19 | 9 | 13 | 1.06 | [0.68; 1.67] | 0.9% | 1.0% |
| Mohan et al | 72 | 80 | 39 | 45 | 1.04 | [0.91; 1.19] | 4.3% | 11.5% |
| Vallejos et al | 139 | 250 | 125 | 251 | 1.12 | [0.95; 1.32] | 10.7% | 7.7% |
| **Fixed effect model** | | **2172** | | **1950** | **1.02** | **[0.97; 1.07]** | **100.0%** | **−−** |
| **Random effects model** | | | | | **1.02** | **[0.97; 1.07]** | **−−** | **100.0%** |

Heterogeneity: $I^2 = 0\%$, $\tau^2 = 0$, $p = 0.89$

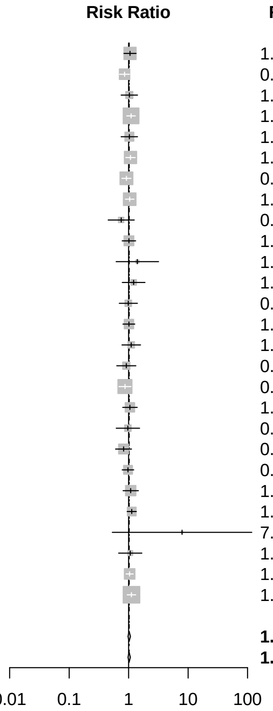


##### Hypertension

| Study | Experimental Events | Experimental Total | Control Events | Control Total | RR | 95%−CI | Weight (fixed) | Weight (random) |
|---|---|---|---|---|---|---|---|---|
| Shouman et al | 16 | 203 | 13 | 101 | 0.61 | [0.31; 1.22] | 5.7% | 4.2% |
| Elgazzar et al − Mild | 11 | 100 | 12 | 100 | 0.92 | [0.42; 1.98] | 3.9% | 3.4% |
| Elgazzar et al − Severe | 14 | 100 | 18 | 100 | 0.78 | [0.41; 1.48] | 5.9% | 4.9% |
| Elgazzar et al − Prophylaxis | 15 | 100 | 14 | 100 | 1.07 | [0.55; 2.10] | 4.6% | 4.5% |
| Krolewiecki et al | 3 | 30 | 3 | 15 | 0.50 | [0.11; 2.19] | 1.3% | 0.9% |
| Chachar et al | 7 | 25 | 6 | 25 | 1.17 | [0.46; 2.98] | 2.0% | 2.3% |
| Babalola et al | 4 | 42 | 5 | 20 | 0.38 | [0.11; 1.27] | 2.2% | 1.4% |
| Kirti et al | 21 | 55 | 18 | 57 | 1.21 | [0.73; 2.01] | 5.8% | 7.9% |
| Okumus et al | 5 | 30 | 8 | 30 | 0.62 | [0.23; 1.69] | 2.6% | 2.0% |
| Beltran et al | 12 | 36 | 14 | 37 | 0.88 | [0.47; 1.64] | 4.5% | 5.3% |
| Lopez−Medina et al | 28 | 200 | 25 | 198 | 1.11 | [0.67; 1.83] | 8.2% | 8.1% |
| Bermejo Galan et al | 21 | 53 | 48 | 115 | 0.95 | [0.64; 1.41] | 9.9% | 12.9% |
| Mahmud et al | 29 | 200 | 28 | 200 | 1.04 | [0.64; 1.68] | 9.2% | 8.8% |
| Abd−Elsalam et al | 18 | 82 | 14 | 82 | 1.29 | [0.69; 2.41] | 4.6% | 5.2% |
| Chahala et al | 13 | 117 | 8 | 117 | 1.62 | [0.70; 3.77] | 2.6% | 2.9% |
| Bukhari et al | 5 | 41 | 7 | 45 | 0.78 | [0.27; 2.28] | 2.2% | 1.8% |
| Seet et al | 8 | 617 | 3 | 619 | 2.68 | [0.71; 10.04] | 1.0% | 1.2% |
| Mohan et al | 9 | 80 | 5 | 45 | 1.01 | [0.36; 2.84] | 2.1% | 1.9% |
| Vallejos et al | 53 | 250 | 66 | 251 | 0.81 | [0.59; 1.11] | 21.6% | 20.3% |
| **Fixed effect model** | | **2361** | | **2257** | **0.95** | **[0.83; 1.10]** | **100.0%** | **−−** |
| **Random effects model** | | | | | **0.95** | **[0.82; 1.09]** | **−−** | **100.0%** |

Heterogeneity: $I^2 = 0\%$, $\tau^2 = 0$, $p = 0.78$

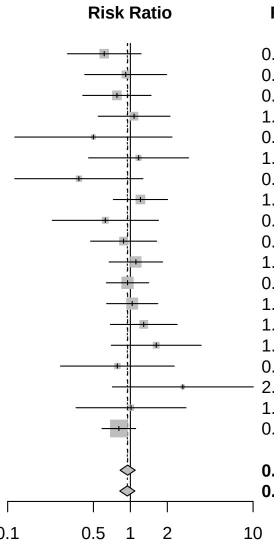


##### Diabetes

| Study | Experimental Events | Experimental Total | Control Events | Control Total | RR | 95%−CI | Weight (fixed) | Weight (random) |
|---|---|---|---|---|---|---|---|---|
| Shouman et al | 13 | 203 | 10 | 101 | 0.65 | [0.29; 1.42] | 5.6% | 4.2% |
| Elgazzar et al − Mild | 15 | 100 | 14 | 100 | 1.07 | [0.55; 2.10] | 5.8% | 5.8% |
| Elgazzar et al − Severe | 18 | 100 | 21 | 100 | 0.86 | [0.49; 1.51] | 8.7% | 8.2% |
| Elgazzar et al − Prophylaxis | 15 | 100 | 19 | 100 | 0.79 | [0.43; 1.46] | 7.9% | 6.8% |
| Krolewiecki et al | 6 | 30 | 1 | 15 | 3.00 | [0.40; 22.71] | 0.6% | 0.6% |
| Chachar et al | 11 | 25 | 8 | 25 | 1.38 | [0.67; 2.83] | 3.3% | 5.0% |
| Babalola et al | 2 | 42 | 0 | 20 | 2.41 | [0.12; 47.97] | 0.3% | 0.3% |
| Kirti et al | 21 | 55 | 19 | 57 | 1.15 | [0.70; 1.88] | 7.8% | 10.5% |
| Okumus et al | 15 | 30 | 12 | 30 | 1.25 | [0.71; 2.20] | 5.0% | 8.2% |
| Beltran et al | 9 | 36 | 16 | 37 | 0.58 | [0.29; 1.14] | 6.6% | 5.7% |
| Lopez−Medina et al | 10 | 200 | 12 | 198 | 0.82 | [0.36; 1.87] | 5.0% | 3.9% |
| Bermejo Galan et al | 14 | 53 | 31 | 115 | 0.98 | [0.57; 1.68] | 8.1% | 8.9% |
| Mahmud et al | 24 | 200 | 29 | 200 | 0.83 | [0.50; 1.37] | 12.1% | 10.3% |
| Abd−Elsalam et al | 17 | 82 | 10 | 82 | 1.70 | [0.83; 3.49] | 4.2% | 5.1% |
| Chahala et al | 10 | 117 | 7 | 117 | 1.43 | [0.56; 3.63] | 2.9% | 3.0% |
| Bukhari et al | 6 | 41 | 4 | 45 | 1.65 | [0.50; 5.42] | 1.6% | 1.8% |
| Seet et al | 2 | 617 | 2 | 619 | 1.00 | [0.14; 7.10] | 0.8% | 0.7% |
| Mohan et al | 6 | 80 | 5 | 45 | 0.68 | [0.22; 2.09] | 2.7% | 2.0% |
| Vallejos et al | 21 | 250 | 27 | 251 | 0.78 | [0.45; 1.34] | 11.2% | 8.9% |
| **Fixed effect model** | | **2361** | | **2257** | **0.97** | **[0.82; 1.14]** | **100.0%** | **--** |
| **Random effects model** | | | | | **0.97** | **[0.83; 1.14]** | **--** | **100.0%** |

Heterogeneity: $I^2 = 0\%$, $\tau^2 = 0$, $p = 0.80$

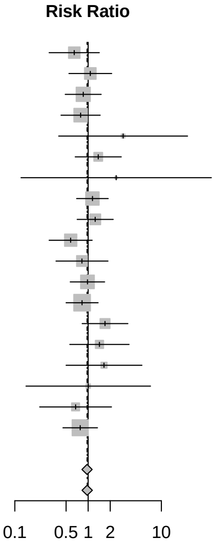


#### Asthma

| Study | Experimental Events | Experimental Total | Control Events | Control Total | RR | 95%−CI | Weight (fixed) | Weight (random) |
|---|---|---|---|---|---|---|---|---|
| Shouman et al | 7 | 203 | 2 | 101 | 1.74 | [0.37; 8.23] | 4.6% | 5.2% |
| Elgazzar et al − Mild | 5 | 100 | 6 | 100 | 0.83 | [0.26; 2.64] | 10.3% | 9.5% |
| Elgazzar et al − Severe | 14 | 100 | 12 | 100 | 1.17 | [0.57; 2.40] | 20.5% | 24.4% |
| Elgazzar et al − Prophylaxis | 5 | 100 | 4 | 100 | 1.25 | [0.35; 4.52] | 6.8% | 7.6% |
| Krolewiecki et al | 4 | 30 | 1 | 15 | 2.00 | [0.24; 16.36] | 2.3% | 2.9% |
| Kirti et al | 1 | 55 | 0 | 57 | 3.11 | [0.13; 74.69] | 0.8% | 1.2% |
| Mahmud et al | 9 | 200 | 12 | 200 | 0.75 | [0.32; 1.74] | 20.5% | 17.8% |
| Vallejos et al | 16 | 250 | 20 | 251 | 0.80 | [0.43; 1.51] | 34.1% | 31.4% |
| **Fixed effect model** | | **1038** | | **924** | **0.99** | **[0.70; 1.41]** | **100.0%** | **--** |
| **Random effects model** | | | | | **0.98** | **[0.69; 1.40]** | **--** | **100.0%** |

Heterogeneity: $I^2 = 0\%$, $\tau^2 = 0$, $p = 0.91$

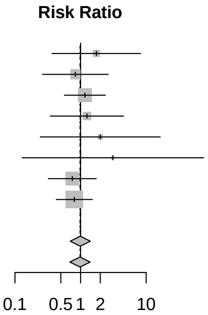


#### Ischemic heart disease

| Study | Experimental Events | Experimental Total | Control Events | Control Total | RR | 95%−CI | Weight (fixed) | Weight (random) |
|---|---|---|---|---|---|---|---|---|
| Shouman et al | 6 | 203 | 1 | 101 | 2.99 | [0.36; 24.46] | 2.9% | 5.6% |
| Elgazzar et al − Mild | 2 | 100 | 6 | 100 | 0.33 | [0.07; 1.61] | 12.9% | 9.9% |
| Elgazzar et al − Severe | 5 | 100 | 12 | 100 | 0.42 | [0.15; 1.14] | 25.8% | 24.4% |
| Elgazzar et al − Prophylaxis | 1 | 100 | 3 | 100 | 0.33 | [0.04; 3.15] | 6.4% | 4.9% |
| Chachar et al | 2 | 25 | 2 | 25 | 1.00 | [0.15; 6.55] | 4.3% | 7.0% |
| Kirti et al | 2 | 55 | 8 | 57 | 0.26 | [0.06; 1.17] | 16.9% | 10.9% |
| Okumus et al | 0 | 30 | 1 | 30 | 0.33 | [0.01; 7.86] | 3.2% | 2.5% |
| Lopez−Medina et al | 4 | 200 | 3 | 198 | 1.32 | [0.30; 5.82] | 6.5% | 11.2% |
| Bukhari et al | 3 | 41 | 2 | 45 | 1.65 | [0.29; 9.37] | 4.1% | 8.2% |
| Mohan et al | 0 | 80 | 1 | 45 | 0.19 | [0.01; 4.53] | 4.1% | 2.4% |
| Vallejos et al | 3 | 250 | 6 | 251 | 0.50 | [0.13; 1.99] | 12.9% | 13.1% |
| **Fixed effect model** | | **1184** | | **1052** | **0.58** | **[0.36; 0.93]** | **100.0%** | **--** |
| **Random effects model** | | | | | **0.58** | **[0.35; 0.95]** | **--** | **100.0%** |

Heterogeneity: $I^2 = 0\%$, $\tau^2 = 0$, $p = 0.62$

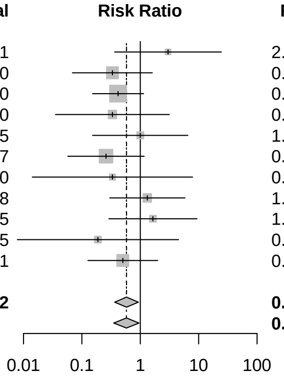


#### Chronic kidney disease

| Study | Experimental Events | Experimental Total | Control Events | Control Total | RR | 95%−CI | Weight (fixed) | Weight (random) |
|---|---|---|---|---|---|---|---|---|
| Shouman et al | 2 | 203 | 1 | 101 | 1.00 | [0.09; 10.84] | 10.6% | 10.8% |
| Kirti et al | 1 | 55 | 2 | 57 | 0.52 | [0.05; 5.55] | 15.7% | 11.0% |
| Beltran et al | 2 | 36 | 1 | 37 | 2.06 | [0.19; 21.69] | 7.9% | 11.1% |
| Bermejo Galan et al | 2 | 53 | 2 | 115 | 2.17 | [0.31; 14.99] | 10.1% | 16.6% |
| Mahmud et al | 3 | 200 | 5 | 200 | 0.60 | [0.15; 2.48] | 39.8% | 30.8% |
| Chahala et al | 3 | 117 | 2 | 117 | 1.50 | [0.26; 8.81] | 15.9% | 19.7% |
| **Fixed effect model** | | **664** | | **627** | **1.04** | **[0.49; 2.23]** | **100.0%** | **--** |
| **Random effects model** | | | | | **1.06** | **[0.48; 2.33]** | **--** | **100.0%** |

Heterogeneity: $I^2 = 0\%$, $\tau^2 = 0$, $p = 0.86$

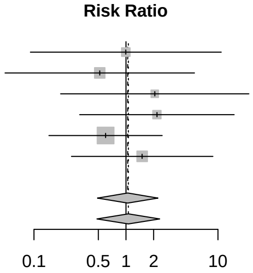

### Obesity

| Study | Experimental Events | Experimental Total | Control Events | Control Total | RR | 95%−CI | Weight (fixed) | Weight (random) |
|---|---|---|---|---|---|---|---|---|
| Krolewiecki et al | 19 | 30 | 12 | 15 | 0.79 | [0.55; 1.15] | 15.6% | 32.7% |
| Chachar et al | 2 | 25 | 4 | 25 | 0.50 | [0.10; 2.49] | 3.9% | 2.0% |
| Lopez−Medina et al | 37 | 200 | 38 | 196 | 0.95 | [0.63; 1.43] | 37.5% | 27.7% |
| Bermejo Galan et al | 22 | 53 | 41 | 115 | 1.16 | [0.78; 1.74] | 25.3% | 28.3% |
| Chahala et al | 10 | 117 | 18 | 117 | 0.56 | [0.27; 1.15] | 17.6% | 9.3% |
| **Fixed effect model** | | **425** | | **468** | **0.89** | **[0.71; 1.13]** | **100.0%** | **−−** |
| **Random effects model** | | | | | **0.89** | **[0.71; 1.12]** | **−−** | **100.0%** |

Heterogeneity: $I^2 = 7\%$, $\tau^2 = 0.0049$, $p = 0.37$

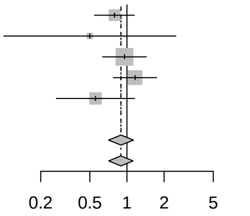


### Chronic pulmonary disease

| Study | Experimental Events | Experimental Total | Control Events | Control Total | RR | 95%−CI | Weight (fixed) | Weight (random) |
|---|---|---|---|---|---|---|---|---|
| Kirti et al | 1 | 55 | 0 | 57 | 3.11 | [0.13; 74.69] | 2.1% | 3.6% |
| Okumus et al | 0 | 30 | 1 | 30 | 0.33 | [0.01; 7.86] | 6.4% | 3.7% |
| Beltran et al | 2 | 36 | 4 | 37 | 0.51 | [0.10; 2.63] | 16.9% | 13.7% |
| Lopez−Medina et al | 6 | 200 | 6 | 198 | 0.99 | [0.32; 3.02] | 25.8% | 29.4% |
| Bermejo Galan et al | 2 | 53 | 7 | 115 | 0.62 | [0.13; 2.88] | 18.9% | 15.4% |
| Vallejos et al | 7 | 250 | 7 | 251 | 1.00 | [0.36; 2.82] | 29.9% | 34.2% |
| **Fixed effect model** | | **624** | | **688** | **0.85** | **[0.47; 1.53]** | **100.0%** | **−−** |
| **Random effects model** | | | | | **0.85** | **[0.46; 1.55]** | **−−** | **100.0%** |

Heterogeneity: $I^2 = 0\%$, $\tau^2 = 0$, $p = 0.89$

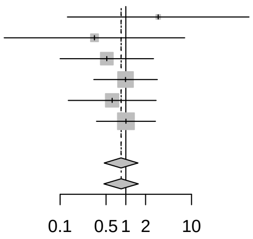


### Cancer

| Study | Experimental Events | Experimental Total | Control Events | Control Total | RR | 95%−CI | Weight (fixed) | Weight (random) |
|---|---|---|---|---|---|---|---|---|
| Kirti et al | 2 | 55 | 4 | 57 | 0.52 | [0.10; 2.72] | 42.2% | 32.0% |
| Okumus et al | 0 | 30 | 1 | 30 | 0.33 | [0.01; 7.86] | 16.1% | 8.8% |
| Bermejo Galan et al | 2 | 53 | 3 | 115 | 1.45 | [0.25; 8.40] | 20.3% | 28.4% |
| Vallejos et al | 4 | 250 | 2 | 251 | 2.01 | [0.37; 10.86] | 21.4% | 30.8% |
| **Fixed effect model** | | **388** | | **453** | **1.00** | **[0.41; 2.42]** | **100.0%** | **−−** |
| **Random effects model** | | | | | **1.01** | **[0.40; 2.59]** | **−−** | **100.0%** |

Heterogeneity: $I^2 = 0\%$, $\tau^2 = 0$, $p = 0.59$

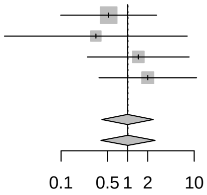
