## Supplemental Data 1 for "Bias as a source of inconsistency in ivermectin trials for COVID-19: A systematic review"

**S2 Table. Included trials' methodological limitations**

| Study | Methodological limitations |
|---|---|
| Shouman et al | There is no information on the randomization process and no concealment of allocation strategy is described, also there is not enough information to assess significant baseline differences between arms. Non-blinded study which may have resulted in deviation from intended interventions and bias in outcome measurement. |
| Chowdhury et al | Study was registered as a non-randomized single center study. Results to be submitted ClinicalTrials.gov by the sponsor or investigator but not yet posted, pending quality control review for apparent errors, deficiencies, or inconsistencies. Quasi-randomisation based on odd-even allocation suggests no allocation concealment. Non-blinded study which may have resulted in deviations from intended interventions and bias in outcome measurements. |
| Podder et al | Quasi-randomisation based on odd-even allocation with no allocation concealment. Non-blinded study which may have resulted in deviations from intended interventions and bias in outcome measurement. |
| Hashim et al | Quasi-randomisation based on odd-even allocation with no allocation concealment. Patients and investigators not blinded which may have resulted in deviations from intended interventions. Outcome assessors also probably not blinded. |
| Elgazzar et al | There is no information on randomization process and no concealment of allocation strategy is described. Not enough information to assess significant baseline differences between arms. The study is presented as "doble-blind" but there is no specification on how this was achieved (e.g., use of placebo), which may have resulted in deviations from intended interventions and bias in the measurement of the outcomes. It is unclear if an appropriate analytical approximation was used as those with clinical failure were shifted to an alternative management protocol with no further specifications. |
| Krolewiecki et al | Non-blinded study which may have resulted in deviation from intended interventions and bias in the measurement of some of the outcomes. |
| Niaee et al | Randomization sequence is described as adequately concealed but baseline differences between intervention groups suggest a problem with the randomization process. Important discrepancies in PCR positivity rates at baseline (47% in placebo, 60% in SOC, and 97% in Arm/Gp 3.) In addition, there is no explanation on why two control arms were implemented (one with placebo and the other without placebo). The study described that clinicians and patients were blinded but is not clear how this was achieved considering that in one of the control arms no placebo was used. |
| Ahmed et al | No concealment of the allocation strategy is mentioned, also there is not enough information to assess significant baseline differences between arms. Unclear whether the |

| | |
|---|---|
| | results were selected from multiple outcome measurements or analyses of the data and/or if the trial was analyzed as pre-specified. |
| Chaccour et al | No significant limitations identified. |
| Chachar et al | No concealment of the allocation strategy is mentioned. Non-blinded study which may result in deviation from intended interventions and bias in the measurement of the outcomes. |
| Babalola et al | The study was reported as “double blind” but lopinavir/ritonavir was not allowed for patients assigned to ivermectin arms. This strategy may have compromised the blinding of physicians and study personnel resulting in deviation from intended interventions and bias in the measurement of the outcomes. |
| Kirti R et al | No significant limitations. |
| Chahla et al | There is no information on randomization process and no concealment of allocation strategy is described, also there is not enough information to assess significant baseline differences between arms. Non-blinded study which may result in deviation from intended interventions and bias in the measurement of the outcomes. |
| Mohan et al | No significant limitations. |
| Shahbaznejad et al | No significant limitations. |
| Samaha et al | No concealment of the allocation strategy is mentioned. Non-blinded study which may have resulted in deviation from intended interventions and bias in the measurement of the outcomes. |
| Bukhari et al | There is no information on randomization process and no concealment of allocation strategy is described, also there is not enough information to assess significant baseline differences between arms. Non-blinded study which may have resulted in deviation from intended interventions and bias in the measurement of the outcomes. |
| Okumus et al | No concealment of the allocation strategy is mentioned and baseline differences between intervention groups suggest a problem with the randomization process.<br>The study was described in the protocol as “non-blinded” study which may have resulted in deviations from intended interventions and bias in the measurement of the outcomes. |
| Beltran et al | Randomization process not described, and no concealment of the allocation strategy is mentioned, however baseline characteristics were apparently balanced. In addition, appropriate blinding strategy is described. |
| Lopez-Medina et al | 38 patients in intervention arm and 40 patients in control arm were excluded after randomization because of a labeling error. We assume that the chance of being excluded was not related to patient’s prognosis hence prognostic balance was probably not affected. |
| Bermejo Galan et al | No significant limitations. |
| Pott-Junior et al | Non-blinded study which may have resulted in deviation from intended interventions and bias in the measurement of the outcomes. |

| | |
|---|---|
| Kishoria et al | Non-blinded study which may have resulted in deviation from intended interventions and bias in the measurement of the outcomes. |
| Seet et al | It is not clear if those in charge of assigning recruited patients to each floor/intervention were aware of the allocation sequence. Non-blinded study which may have resulted in deviation from intended interventions and bias in the measurement of outcomes. |
| Mahmud et al | Overall, 9.3% on randomized patients excluded from analysis. 15 participants lost to follow-up in the intervention and 17 participants in the control arm; 3 participants that died in the control group and 2 in the intervention group due to adverse events, were also excluded for viral clearance outcome. There is no evidence that the result was not biased by missing outcome data. |
| Abd-Elsalam et al | Non-blinded study which may have resulted in deviation from intended interventions and bias in the measurement of the outcomes. |
| Biber et al | The allocation sequence was described as adequately concealed but not enough information available on baseline differences between intervention groups. 7 (10.4%) of patients excluded in the intervention arm vs. 14 (18.4%) in control arm because RT-PCR with Ct value >35 which suggests potential baseline imbalances between arms. In addition, 3 patients were lost to follow up in each arm. |
| Faisal el al | There is no information on randomization process and no concealment of allocation strategy is described, also there is not enough information to assess significant baseline differences between arms. Non-blinded study which may have resulted in deviation from intended interventions and bias in the measurement of the outcomes. |
| Vallejos et al | Although allocation concealment strategy was not appropriately described in the manuscript, authors provided further details by email. Appropriate blinding strategy was described. |
